# Predicting the onset of internalizing disorders in early adolescence using deep learning optimized with AI

**DOI:** 10.1101/2023.08.21.23294377

**Authors:** Nina de Lacy, Michael J. Ramshaw

## Abstract

Internalizing disorders (depression, anxiety, somatic symptom disorder) are among the most common mental health conditions that can substantially reduce daily life function. Early adolescence is an important developmental stage for the increase in prevalence of internalizing disorders and understanding specific factors that predict their onset may be germane to intervention and prevention strategies. We analyzed ∼6,000 candidate predictors from multiple knowledge domains (cognitive, psychosocial, neural, biological) contributed by children of late elementary school age (9-10 yrs) and their parents in the ABCD cohort to construct individual-level models predicting the later (11-12 yrs) onset of depression, anxiety and somatic symptom disorder using deep learning with artificial neural networks. Deep learning was guided by an evolutionary algorithm that jointly performed optimization across hyperparameters and automated feature selection, allowing more candidate predictors and a wider variety of predictor types to be analyzed than the largest previous comparable machine learning studies. We found that the future onset of internalizing disorders could be robustly predicted in early adolescence with AUROCs ≥∼0.90 and ≥∼80% accuracy. Each disorder had a specific set of predictors, though parent problem behavioral traits and sleep disturbances represented cross-cutting themes. Additional computational experiments revealed that psychosocial predictors were more important to predicting early adolescent internalizing disorders than cognitive, neural or biological factors and generated models with better performance. We also observed that the accuracy of individual-level models was highly correlated to the relative importance of their constituent predictors, suggesting that principled searches for predictors with higher importance or effect sizes could support the construction of more accurate individual-level models of internalizing disorders. Future work, including replication in additional datasets, will help test the generalizability of our findings and explore their application to other stages in human development and mental health conditions.

## INTRODUCTION

Depression, anxiety and problematic somatic symptoms (physical symptoms such as headaches and stomachaches) are common mental health issues in adolescence. Often collectively referred to as internalizing disorders, they have been associated with reduced levels of well-being and daily life function, increased risk of self-harm and suicide and are substantial predictors of adult psychopathology. (1) Depression and anxiety are among the most common mental illnesses in the population with lifetime prevalence of ∼30% and ∼20% respectively. (2) The incidence of internalizing disorders increases exponentially during the peri-adolescent period, with anxiety having an earlier developmental arc. (3) Anxiety disorders emerge during elementary school, with the median age of onset being 11 years of age (yrs) and 75% of lifetime illness occurring by 21 yrs. Major depression cases begin to onset at 11-12 yrs with median onset at 31-32 yrs and 75% of lifetime illness having onset by 44 yrs. (4) Problematic somatic symptoms affect up to 40% of youth and increase over peri-adolescence: one third to a half continue to report symptoms as adults with 5-7% in the general population and ∼17% in the primary care population meeting criteria as adults for Somatic Symptom Disorder (SSD). (5, 6)

Given the considerable personal, societal and economic burdens associated with internalizing disorders, (7, 8, 9, 10) there is great interest in identifying specific factors that predict their onset, since evidence suggests that early intervention improves outcomes (11, 12) and reduces resource use. (13) Isolating key predictors of internalizing disorders is challenging since they have been associated with a host of different factors from varied domains ranging from biological (neural; genetic; hormonal) and psychological models (fear/threat response) to interpersonal relationship function, parent characteristics, the community environment and wider social determinants of health such as relative poverty. Historically, an important barrier to disambiguating the relative importance of such factors to predicting case onset has been the paucity of appropriate multimodal data in large participant samples. Outside the US, national registries or school system data have been available offering large sample sizes (*n*>10,000) but these typically lack physiologic information such as neuroimaging data. (14, 15, 16, 17) An alternative strategy is to combine data from multiple studies offering neuroimaging or genomic data to boost sample size such as the datasets offered by IMAGEN or ENIGMA, though pooling across heterogenous studies may inherently limit features (variables) available for analysis to those that are shared across all studies. (**18, 19, 20**) Consequently, to promote comparative discovery at scale, federal and other organizations have recently sponsored the formation of large, longitudinal cohorts collecting a wide variety of multimodal data types with standardized protocols. In peri-adolescence, the flagship initiative of this type is the ongoing population-level ABCD study (*n*=11,800) used in the present study. (21, 22, 23)

Concomitantly, interest has recently grown in applying machine learning (ML) methods to these newly-emerging large-scale population cohorts as ML techniques offer advantages in approaching such high-dimension data. Firstly, they can generate individual-level case predictions from multidimensional data to bridge extant work focused on group-level statistical effects with individual-level discoveries of potential clinical relevance by “providing multivariate signatures that are valid at the single-subject level”. (**24, 25**) Secondly, ML techniques can simultaneously analyze hundreds of candidate predictors and incorporate non-linear relationships among a set of predictors. These properties are relevant to the construction of individual-level models since significant group-level effects may not be useful at the individual level while a feature with low effect size at the group level may prove germane. While a number of ML predictive studies have been performed in youth internalizing disorders, these have to date considered <200 candidate predictors and focused largely on prevailing cases of depression, rather than new onset cases in adolescence, especially early adolescence. The latter are of considerable translational interest since understanding individual-level drivers of illness onset and obtaining better visibility into whether future onset can be reliably predicted using ML would potentially inform intervention strategies. Extant work is also highly heterogenous with respect to which candidate predictors (input features) are considered. In particular, some studies use only psychosocial features and some only neuroimaging features, while a few have incorporated both types. Concomitantly, performance has been variable, with accuracy ranging over ∼50-90% but the achievement of robust precision (positive predictive value) - an important metric for translational relevance - typically proving more elusive. Moreover, since obtaining physiologic measures such as neuroimaging metrics is complex and uncommon in clinical practice, it is relevant to understand whether they improve individual-level case prediction. Finally, few studies have constructed predictive models of anxiety or somatic problems in youth using ML classifiers or applied a consistent analytic architecture across the three major categories of internalizing disorders simultaneously in the same population and data to enable direct comparisons and determine the specificity of predictive models to different internalizing disorders.

In the present study, we aim to build on prior work by predicting cases of depression, anxiety and SSD in early adolescence (9-12 yrs) using deep learning guided by a large-scale AI optimization process. Specifically, we aimed to a) identify and rank the most important predictors after analyzing thousands of multidomain candidate predictors; b) provide individual-level predictions of future, new onset cases at 11-12 yrs in comparison to all prevailing cases at the same age and 9-10 yrs; c) determine the incremental value of using multidomain predictors vs neural-only modeling; and d) examine the relationship between predictor importance and accuracy. Applying a common analytic architecture to data from the ABCD cohort, we first constructed multimodal predictive models by analyzing 5,777 candidate predictors spanning demographics; developmental and medical history; white and gray matter brain structure, neural function (cortical and subcortical connectivity, 3 tasks); brain volumetrics; physiologic function (e.g. sleep, hormone levels, pubertal stage, physical function); cognitive and academic performance; social and cultural environment (e.g. parents, friends, bullying); activities of everyday life (e.g. screen use, hobbies); living environment (e.g. crime, pollution, educational and food availability) and substance use. Subsequently, we recapitulated all analytic procedures using multiple types of neural candidate predictors.

To make these case classifications, we used deep learning with artificial neural networks, which incorporates non-linear relationships among predictors and is resistant to multicollinearity. While artificial neural networks offer powerful predictive capability, their application to translational aims can be limited by the relative difficulty of tuning these models (setting hyperparameters that control learning) and their tendency to act as ‘black box’ estimators where the features used to make predictions are not interpretable and their relative importance is difficult to determine. We enhanced deep learning performance with Integrated Evolutionary Learning (IEL), an AI-based form of computational intelligence, to jointly optimize across the hyperparameters and learn the most important final predictors and render interpretable predictions. IEL is a genetic algorithm which instantiates the principles of natural selection in computer code, typically performing ∼40,000 model fits during training before testing final, optimized models in a holdout, unseen data partition. All results presented are from testing for generalization in this holdout, unseen data.

## MATERIALS AND METHODS

### Terminology and definitions

Terms used in quantitative analysis may be shared among different fields with variant meanings. Here, we use ML conventions throughout. (26, 27, 28) ‘Prediction’ means predicting the quantitative value of a target variable by analyzing patterns in input data. We refer to the set of all input data as containing ‘features’ or ‘candidate predictors’ and those identified in final, optimized models (presented in **Results)** as ‘final predictors’. The set of observations used to train and validate models is referred to as the ‘training set’ and the unseen holdout set of observations is termed the ‘test set’. We use ‘generalizability’ to refer to the ability of a trained model to adapt to new, previously unseen data drawn from the same distribution i.e. model fit in the test set. ‘Precision’ refers to the fraction of positive predictions that were correct; ‘Recall’ to the proportion of true positives that were correctly predicted; and ‘Accuracy’ to the number of correct predictions as a fraction of total predictions. Receiver Operating Characteristic curves (ROC Curves) are provided that quantify classification performance at different classification thresholds plotting true positive versus false positive rates, where the Area Under the Curve (AUROC) is defined as the two-dimensional area under the ROC curve from (0,0) to (1,1).

### Data and data collection in the ABCD study

Data used in the present study comes from the ABCD study, an epidemiologically informed prospective cohort study that is the largest study of brain development and child health conducted in the United States to date. ABCD recruited 11,880 children (52% male; 48% female) at ages 9-10 years (108-120 months) via 21 sites across the United States and will follow this cohort until age 19-20. The cohort is oversampled for twin pairs (*n*=800) and non-twin siblings from the same family may also be enrolled. A wide variety of information is collected about participants. This data has been made available to qualified researchers at no cost from the National Institute of Mental Health Data Archive since 2018 and is released periodically. This study uses data from release 4.0, which includes data up to the 42-month follow-up date. A full explanation of recruitment procedures, the participant sample and overall design of the ABCD study may be found in Jernigan et al; Garavan et al; and Volkow et al. (29, 30, 31) This study has been reviewed and deemed not human subjects research by the University of Utah Institutional Review Board.

The phenotypic and substance abuse assessment protocol is covered in detail in Barch et al and Lisdahl et al, respectively. (32, 33) In brief, phenotypic assessments of physical and mental health, substance use, neurocognition and culture and environment are performed for youth and their parents and biospecimen collection for DNA, pubertal hormone levels, substance use metabolites (hair) and substance and environmental toxin exposure (baby teeth) are collected from youth at 9-10 yrs. A summary description of assessments performed and environmental and school-related variables derived from geocoding at age 9-10 yrs surveyed in the present study may be inspected in **Supplementary Table 1**.

Brain imaging is collected at 9-10 yrs and every two years thereafter and incorporates optimized 3D T1; 3D T2; Diffusion Tensor Imaging; Resting state functional MRI (rsfMRI); and 3 task MRI (tfMRI) protocols that are harmonized to be compatible across acquisition sites. The tfMRI protocol comprises the Monetary Incentive Delay (MID) and Stop Signal (SST) tasks and an emotional version of the n-back task which collectively measure reward processing, motivation, impulsivity, impulse control, working memory and emotion regulation. The ABCD study provides fully-processed metrics from each of these imaging types. Full details of the neuroimaging protocol may be inspected in Casey et al and the pre-processing and analytic pipeline used to generate neural metrics in Hagler et al. (34, 35) The present study uses all available processed metrics that have passed quality control from the diffusion fullshell; cortical and subcortical Gordon correlations (derived from rsfMRI); structural; volumetric; and all three tasks as well as corresponding head motion statistics for each modality. For certain modalities such as rsfMRI, multiple scans were attempted or completed. In such cases we use variables from the first scan.

### Study inclusion criteria and sample partitioning for machine learning

Inclusion criteria for the present study were a) participants enrolled in the study at baseline who were still enrolled at 2-year follow-up (*n*=8,085) who had b) complete data passing quality control available for all neural metric types (*n*=6,178) and were c) youth participants unrelated to any other youth participant in the study (*n*=5,355). If a youth had a twin or other sibling(s) present in the cohort, we selected the older or oldest sibling for inclusion in our study. We present characteristics of the study sample at 9-10 yrs since these participants correspond to the input data used to make predictions. Demographic characteristics of this sample at age 9-10 yrs are presented in **Table 1**.

**Table 1:** Demographic characteristics of participant sample at age 9-10 years. Sex refers to sex assigned at birth on the original birth certificate. Gender refers to the youth’s gender identification. Race and ethnicity refer to the parents’ view of youth’s race or ethnicity. More than one race or ethnicity identification may be selected and therefore percentages sum to >100%.

| Characteristic | Number | Percent |
| --- | --- | --- |
| **Sex** |  |  |
| Male | 2,771 | 51.7% |
| Female | 2,584 | 48.3 |
| **Gender Identity** |  |  |
| Male | 2,768 | 51.7% |
| Female | 2,577 | 48.1 |
| Gender non-conforming | 7 | 0.1 |
| Don't know/didn't answer | 4 | 0.1 |
| **Race** |  |  |
| Black/African American | 873 | 16.3% |
| Asian | 353 | 6.6 |
| White | 4,236 | 79.1 |
| Native American/Alaska Native | 187 | 3.5 |
| Other | 334 | 6.2 |
| **Ethnicity** |  |  |
| Hispanic/Latino/Latinx | 1,070 | 20.0% |
| Non-Hispanic | 4,224 | 78.9 |
| Not indicated | 62 | 1.2 |

Physiologic and cognitive characteristics of the participant sample at 9-10 yrs may be viewed in **Table 2**.

**Table 2:**
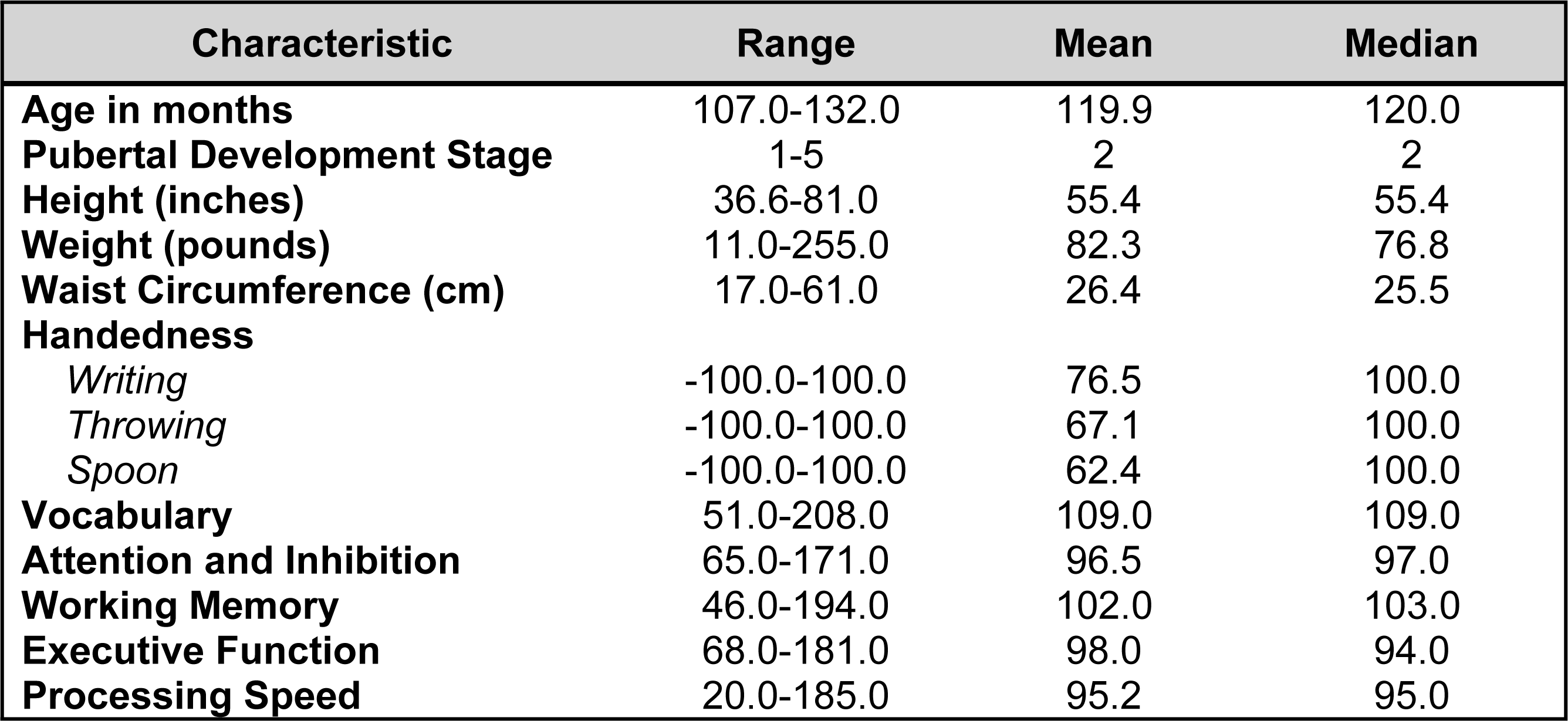
Physiologic and cognitive characteristics of participant sample at age 9-10 years. Characteristics of the study sample at 9-10 yrs. Pubertal development is measured with the Pubertal Development Scale (adapted from the Petersen scale) in a sex-specific manner. Height is measured twice with the average of these values presented. We note a range of 11.0-255.0 pounds for weight which is the range present in the original ABCD data. is assessed with the Edinburgh Handedness Inventory. Cognitive metrics are assessed with the NIH Toolbox and are all age-corrected scores. Vocabulary is measured with the Picture Vocabulary test; Attention and inhibition with the Flanker Inhibitory Control & Attention Task; Executive Function with the Dimensional Change Card Sort Test; and Processing Speed with the Pattern Comparison Processing Speed Test.

| Characteristic | Range | Mean | Median |
| --- | --- | --- | --- |
| Age in months | 107.0-132.0 | 119.9 | 120.0 |
| Pubertal Development Stage | 1-5 | 2 | 2 |
| Height (inches) | 36.6-81.0 | 55.4 | 55.4 |
| Weight (pounds) | 11.0-255.0 | 82.3 | 76.8 |
| Waist Circumference (cm) | 17.0-61.0 | 26.4 | 25.5 |
| Handedness |  |  |  |
| Writing | -100.0-100.0 | 76.5 | 100.0 |
| Throwing | -100.0-100.0 | 67.1 | 100.0 |
| Spoon | -100.0-100.0 | 62.4 | 100.0 |
| Vocabulary | 51.0-208.0 | 109.0 | 109.0 |
| Attention and Inhibition | 65.0-171.0 | 96.5 | 97.0 |
| Working Memory | 46.0-194.0 | 102.0 | 103.0 |
| Executive Function | 68.0-181.0 | 98.0 | 94.0 |
| Processing Speed | 20.0-185.0 | 95.2 | 95.0 |

The resulting group of 5,356 participants was then randomly partitioned into a training set comprising 70% of the sample (*n*=3,749) and a holdout, unseen test set comprising 30% of the sample (*n*=1,607, **Figure 1**). This partitioning was performed prior to pre-processing either features or predictive target to minimize bias.

**Figure 1:**
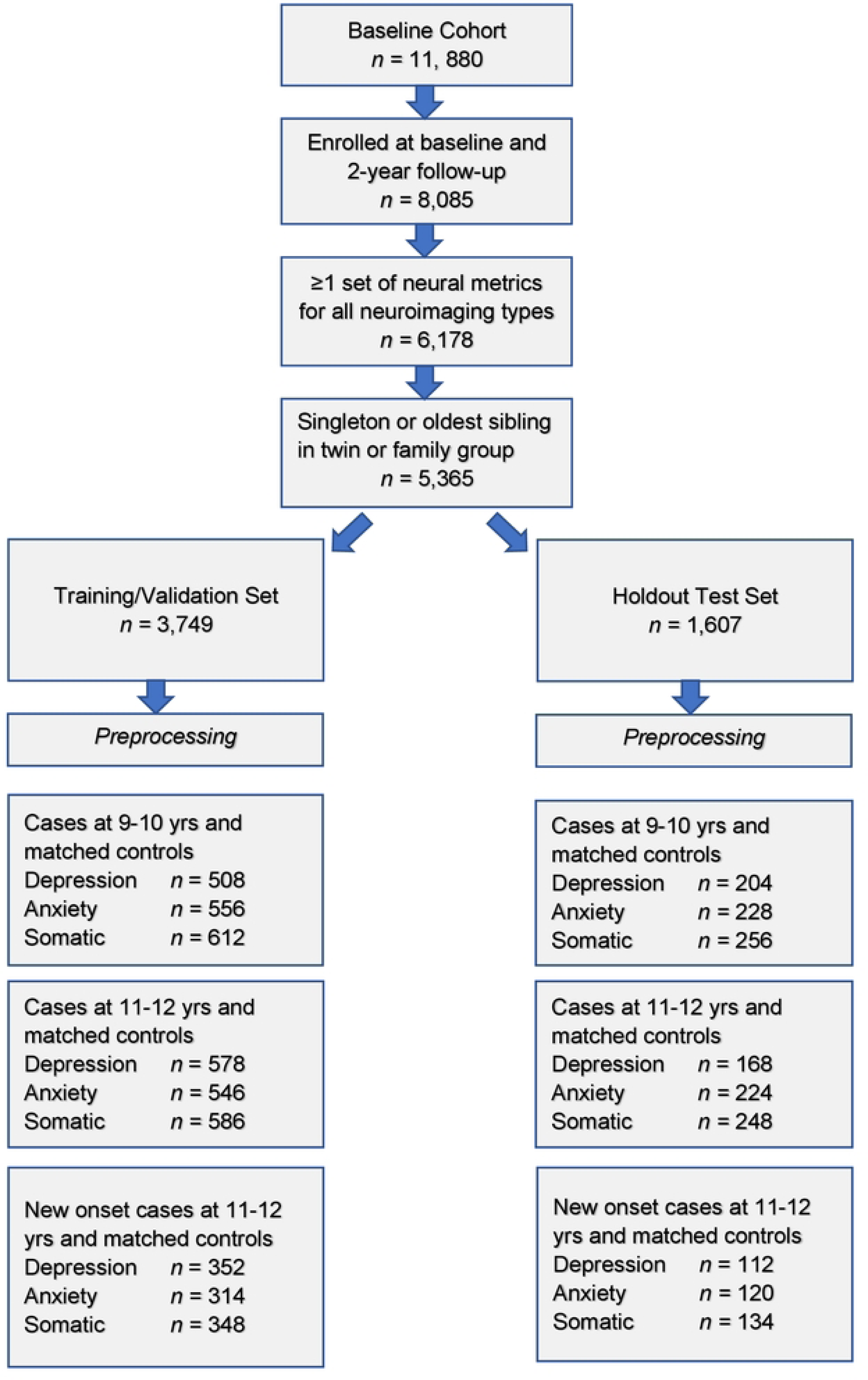
Formation of the study participant sample for internalizing disorders. Steps in the formation of the study sample used to construct predictive models of depression, anxiety and somatic symptom disorder are shown. After exclusion criteria are applied, the sample was randomly partitioned into training and test sets followed by separate pre-processing of targets and features. Subsequently, samples for each experiment were formed as described in **Preparation of predictive targets** and **Construction of participant case samples for internalizing disorders and controls**.

### Preparation of predictive targets

The present study uses predictive targets of depression, anxiety and somatic problems derived from the Child Behavior Checklist for youth ages 4-18 years (CBCL) called the ‘ABCD Parent Child Behavior Checklist Scores Aseba (CBCL) in the ABCD study. The CBCL is a standardized instrument in widespread clinical and research use for the assessment of mental and emotional well-being in youth. It forms part of the Achenbach System of Empirically Based Assessment (ASEBA) “designed to facilitate assessment, intervention planning and outcome evaluation among school, mental health, medical and social service practitioners who deal with maladaptive behavior in children, adolescents and young adults.” (36) During assessment with the CBCL, parents rate their child on a 0-1-2 scale on 118 specific problem items such as “Unhappy, sad or depressed” or “Acts too young for age” for the prior 6 months. The answers to these questions are aggregated into raw, T and percentile scores for 8 syndrome subscales (Anxiety, Somatic Problems, Depression, Social Problems, Thought Problems, Attention Problems, Rule Breaking and Aggressive Behavior) derived from principal components analysis of data from 4455 children referred for mental health services. The CBCL is normed in a sex/gender-specific manner on a U.S. nationally representative sample of 2368 youth ages 4-18 that takes into account differences in problem scores for “males versus females”. It exhibits excellent test-retest reliability of 0.82-0.96 for the syndrome scales with an average *r* of 0.89 across all scales. Content and criterion validity is strong with referred versus non-referred children scoring higher on 113/188 problem items and significantly higher on all problem scales, respectively.

To form binary classification targets for prediction, we thresholded CBCL subscale T scores for Depression, Anxiety and Somatic problems using cutpoints established by ASEBA for clinical practice. Specifically, a T score of 65-69 (95^th^ to 98^th^ percentile) is considered in the ‘borderline clinical’ range, and scores of ≥70 are considered in the ‘clinical range.’ Accordingly, we discretized T scores for each of the 3 subscales under consideration by deeming every individual with a T score ≥ 65 as a ‘case’ [1] and every individual with a score <65 as a ‘not case’ [0]. This process was performed separately for CBCL scores at baseline and 2-year follow-up in the training and test sets.

### Construction of participant case samples for internalizing disorders and controls

To test our hypotheses, we formed 3 different participant samples for each of the internalizing disorders in the training and test sets, respectively (**Figure 1**). The first sample contained cases of depression, anxiety and SSD as defined in **Preparation of predictive targets** at baseline assessment, when youth were 9-10 years of age. The second sample contained cases of depression, anxiety and SSD at 2-year follow-up, when youth were 11-12 years of age. Finally, the third sample contained only new onset cases of depression, anxiety and SSD at 2-year follow-up. A new onset case was defined as a youth who met criteria for depression, anxiety or SSD following the ASEBA threshold in the CBCL who did not meet criteria for the disorder in question at baseline assessment. In all samples, we constructed a balanced sample of controls matched for age and sex/gender selected from the eligible study population (see: **Baseline inclusion criteria and sample partitioning for machine learning** above) from youth with the lowest possible scores on the relevant syndrome scale. No sample in the training sets was <200 participants, a recommended threshold for robust ML analyses.

### Preparation of candidate predictors (input features)

The feature set in the present study comprises the majority of available phenotypic and environmental variables derived from baseline assessment at 9-10 years of age (including data collection site) and all available neural metrics (including head motion statistics) with the exception of temporal variance measures. For continuous phenotypic features where subscale or total scores for assessments were available, these were used. For example, subscale scores for different types of sleep-related disorders from the larger Munich Chronotype Questionnaire. Any metrics or instruments that directly quantified mental health symptoms were excluded since we aimed to predict cases of mental illness without using symptoms. For example, the Youth 7UP Mania scale. The feature set was then partitioned into training and test sets that conformed with the partitions detailed above in **Formation of the study participant sample for internalizing disorders** in **Figure 1**. Pre-processing of phenotypic and environmental features was subsequently performed separately in the training and test sets. First, features with >35% missing values were discarded. This threshold was used since prior research shows that good results may be obtained with ML methods with imputation up to 50% missing data. (**37**) Nominal variables were one-hot encoded to transform them into discrete variables. Continuous variables were then trimmed to [mean +/- 3] standard deviations to remove outliers and all features scaled in the interval [0,1] with the MinMaxScaler. Missing values were imputed using non-negative matrix factorization (NNMF). NNMF is a mathematically-proven imputation method that minimizes the cost function of missing data rather than assuming zero values. It is effective at capturing both global and local structure in the data and it has been demonstrated to perform well regardless of the underlying pattern of missingness. (38, 39, 40) **Supplementary Table 2** shows the number and percentage of observations which were trimmed and filled with NNMF for the training and test sets, respectively. After imputation with NNMF, any variables originating from phenotypic assessments lacking summary scores were reduced to a summary metric using feature agglomeration to produce a final set of (*n=*763) phenotypic and environmental features. Neural metrics (*n*=5,014) were processed and underwent quality control by the ABCD study team and were therefore not pre-processed with the exception of scaling, again performed separately in the training and test partitions. There were no missing neural features. The final combined feature set including neural, phenotypic, environmental, head motion and site features comprised 5,777 features.

### Overview of predictive analytic pipeline

We used deep learning with artificial neural networks (AdamW optimizer) to predict cases of depression, anxiety and somatic problems in early adolescence in three scenarios: at 9-10 years of age, at 11-12 years of age and in new onset cases at 11-12 years of age. Deep learning models were implemented with *k*-fold cross-validation and trained by an AI meta-learning algorithm that jointly performed feature selection and optimized across the hyperparameters in an automated manner, pursuing ∼40,000 model fits for each experiment. Model training was terminated based on the Bayes Information Criterion (BIC), an information theoretic metric. Subsequently, final optimized models were tested for their ability to generalize in the holdout, unseen test set and performance statistics of AUROC, accuracy, precision and recall, and ROC curves are reported for the best-performing models. We also report the relative importance of final predictors to making case predictions quantified with two techniques: Shapley Additive Explanations (SHAP) and permutation using the eli5 algorithm. Detailed explanations of these methods are provided below. Code for the predictive analytics may be accessed at the de Lacy Laboratory GitHub: https://github.com/delacylab/integrated_evolutionary_learning

### Coarse feature selection

Prior to beginning model training, we performed coarse feature selection for each of the nine experiments i.e. 3 targets of depression, anxiety and SSD each in 3 participant samples of 9-10 yrs; 11-12 yrs and new onset cases at 11-12 yrs. The purpose of this process was to quantify, for each sample, which of the 5,777 features exhibited a non-zero relationship with the target in order to reduce the number of features entering the deep learning pipeline in a principled manner. First, a simple filtering process was performed in which χ^2^ (categorical features) and ANOVA (continuous features) statistics and mutual information metric (all features) were computed to quantify the relationship between all features and the target, where the target (depression, anxiety, SSD) was represented by a categorical vector in [0,1]. Any feature with a non-zero relationship (either positive or negative) with the target was retained. Subsequently, feature selection was performed on these filtered feature subsets using the Least Absolute Shrinkage and Selection Operator (LASSO) algorithm. The LASSO is a popular regularization technique based in linear regression that efficiently selects a reduced set of features by forcing certain regression coefficients to zero. The LASSO algorithm has a hyperparameter (commonly called the α) that instantiates the amount of penalization (shrinkage) that will be imposed on the features. We implemented the LASSO with our AI meta-learning algorithm Integrated Evolutionary Learning to tune the α hyperparameter in the same manner as described below in **Integrated Evolutionary Learning for deep learning optimization**.

The number of features retained for each of the 9 experiments after each step in the coarse feature selection process may be examined in **Table 3**. Specific features selected by the LASSO and the resulting univariate coefficients between each of these features and the target vectors (depression, anxiety, somatic problems) for each participant sample (9-10 yrs; 11-12 yrs and new onset cases at 11-12 yrs) may be viewed in **Supplementary Table 3a-i**. Each feature set selected by the LASSO then entered the deep learning pipeline.

**Table 3:** Feature sets after coarse feature selection. The total baseline set of 5,777 features was reduced via coarse feature selection in a two-step process of filtering followed by regularization with the LASSO algorithm. This table displays the number of remaining features after each step for each target (depression, anxiety and somatic problems) and participant sample (at age 9-10 years, at age 11-12 years and for new onset cases at age 11-12 years). Detailed tables showing the univariate coefficients between each feature selected by the LASSO and the target vectors for each case sample and controls may be viewed in **Supplementary Table 3a-i**.

|  | Number of features after filtering | Number of features after selection with LASSO |
| --- | --- | --- |
| **Depression, age 9-10 years** | 4,271 | 133 |
| **Anxiety, age 9 -10 years** | 4,338 | 133 |
| **Somatic problems, age 9-10 years** | 4,357 | 86 |
| **Depression, age 11-12 years** | 4,261 | 35 |
| **Anxiety, age 11-12 years** | 4,295 | 129 |
| **Somatic problems, age 11-12 years** | 4,267 | 215 |
| **Depression, new onset age 11-12 years** | 4,284 | 60 |
| **Anxiety, new onset age 11-12 years** | 4,201 | 85 |
| **Somatic problems, new onset age 11-12 years** | 4,278 | 124 |

### Deep learning with artificial neural networks

We used deep learning to predict cases of depression, anxiety and somatic problems in each participant sample (at ages 9-10, ages 11-12 and for new onset cases only at ages 11-12 years). In order to determine the relative ability of features to predict future cases of internalizing disorders, features collected at baseline assessment (ages 9-10 years) were used to predict cases present at ages 11-12 years. We also constructed similar models that restricted the cases at 11-12 years of age to only new onset cases, where the participant was not exhibiting clinical levels of symptoms at ages 9-10 years. Finally, to quantify any dropoff in predictive power over the two-year followup period, comparative models predicting cases at 9-10 years of age were also computed. Therefore, the feature set comprised only variables collected at 9-10 years of age in all analytic scenarios (**Figure 2**).

**Figure 2:**
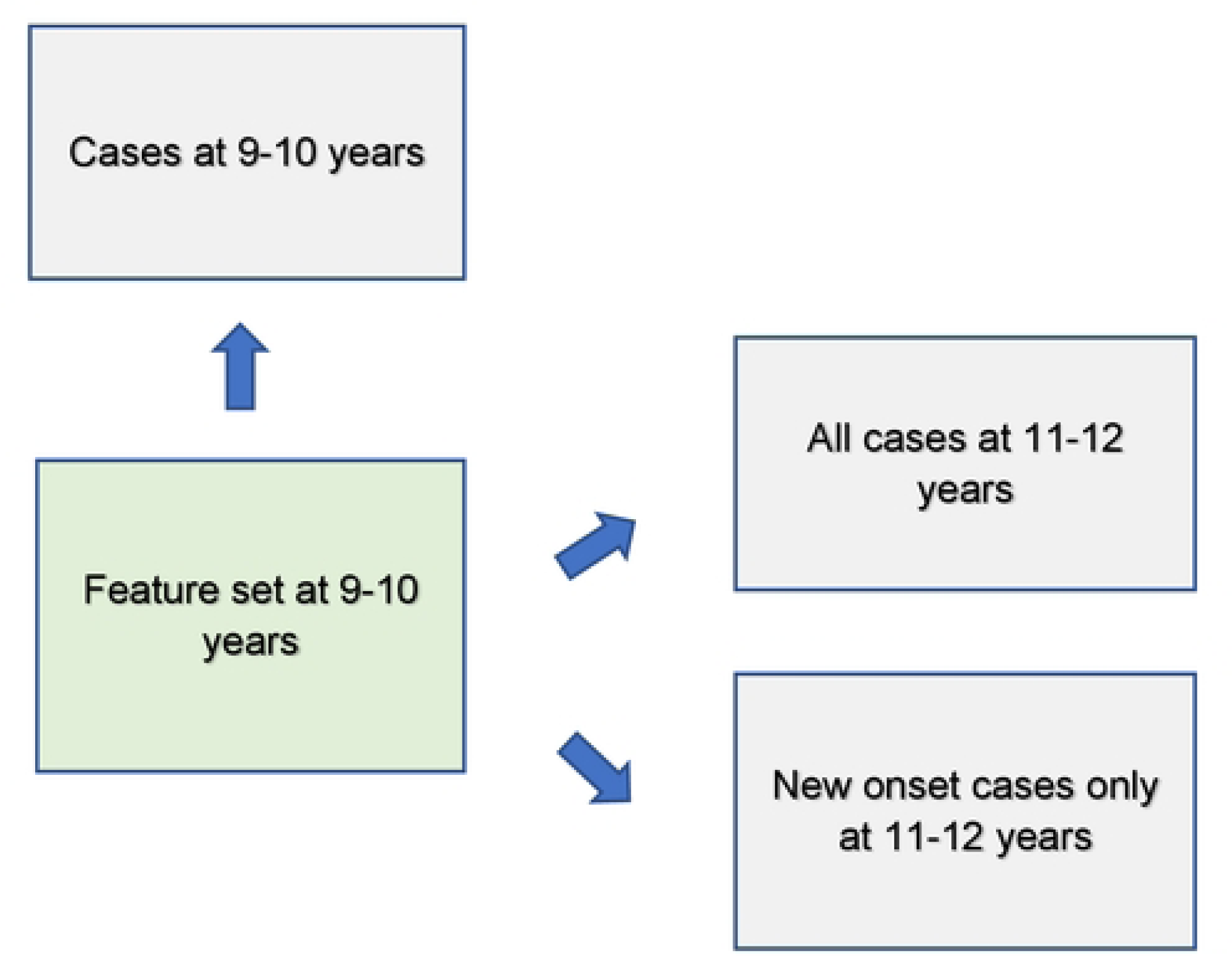
Analytic schema. Features assessed at baseline (ages 9-10 years) were used to predict cases of depression, anxiety and somatic problems present contemporaneously as well as all cases 2 years in the future (ages 11-12 years) and only new onset cases at ages 11-12 years.

We trained artificial neural networks using the AdamW algorithm with 3 layers, 300 neurons per layer, early stopping (patience = 3, metric = validation loss) and the Relu activation function. The last output layer contained a conventional softmax function. Learning parameters (**Table 3**) were tuned with IEL as detailed below. Deep learning models were encoded with TensorFlow embedded in custom Python code.

### Integrated Evolutionary Learning for optimization across hyperparameters and fine feature selection

Many ML algorithms have hyperparameters that control learning. Their settings require ‘tuning’ that can have a dramatic effect on performance. Typically, tuning is performed via ‘rules of thumb’ and ≤50 model fits are explored, introducing the possibility of bias and potentially limiting the solution space. (41, 42, 43) To address this issue, we previously developed and here applied an AI technique called Integrated Evolutionary Learning (IEL) which can improve the performance of ML predictive algorithms in comparable tabular data by up to 20-25% versus the use of default model hyperparameters and conventional designs. (44) IEL is a form of computational intelligence or metaheuristic based on an evolutionary algorithm that instantiates the concepts of biological evolutionary selection in computer code. It optimizes across the hyperparameters of the deep learning algorithm by adaptively breeding models over hundreds of learning generations by selecting for improvements in a fitness function (here, the Bayes Information Criterion, BIC).

For each experiment, the deep learning algorithm was nested inside IEL, which initialized the first generation of 100 models with randomized hyperparameter values or ‘chromosomes’. These hyperparameter settings (**Table 4**) were subsequently recombined, mutated or eliminated over successive generations. In recombination, ‘parent’ hyperparameters were arithmetically averaged to form ‘children’. In mutation, hyperparameter settings were shifted with the range of possible values shown in **Table 4**. When these first 100 models were trained, the BIC was computed for each solution. Of the 80 best models, 40 were recombined by averaging the hyperparameter setting after a pivot point at the midpoint to produce 20 ‘child’ models. 20 were mutated to produce the same number of child models by shifting the requisite hyperparameter by the mutation shift value (**Table 4**). The remaining 20 were discarded. The next generation of models was then formed by adding 60 new models with randomized settings and adding these to the 40 child models retained from the initial generation. Thereafter, IEL continued to recombine, mutate and discard 100 models per generation in a similar fashion to minimize the BIC until the latter fitness function plateaued. With 100 models fitted per generation, IEL typically fits ∼40,000 models per experiment over ∼400 generations.

**Table 4:** Hyperparameter settings optimized with Integrated Evolutionary Learning. Optimization across the hyperparameters of learning rate, Beta 1 and Beta 2 was conducted for deep learning with artificial neural networks within the ranges shown.

| Hyperparameters | Range | Mutation Shift |
| --- | --- | --- |
| Learning rate | 0.00001-0.01 | 0.0001 |
| Beta 1 | 0.9-0.999 | 0.001 |
| Beta 2 | 0.9-0.999 | 0.001 |

IEL jointly performs optimization across hyperparameter settings with automated feature selection and mitigate the risk of overfitting. For each experiment, IEL has available to it the set of features selected in the two-step feature selection process performed with filtering and the LASSO (**Coarse feature selection, Supplementary Table 3**). From each of these sets, a random number of features in the range [2-50] was set for each model in the initial generation of 100 models and specific features were randomly sampled from the set of available features. After computing the BIC for each model, feature sets from the best-performing 60 models were individually allocated to the recombined and mutated child models. Other feature sets were discarded. As with hyperparameter tuning, this process was repeated for succeeding generations until the BIC plateaued.

IEL implements recursive learning to facilitate computationally efficiency. After training until the BIC plateaued, we determine the elbow of the fitness function plotted versus number of features and re-start learning with a warm start. The feature set available after this warm start is constrained to that subset of features, thresholded by their importance, corresponding to the fitness function elbow. Learning then proceeds by thresholding features available for learning at the original warm start feature importance + 2 standard deviations. In addition, the number of models per generation is reduced to 50 and 20 models are recombined and 10 models are mutated. Otherwise, training after the warm start uses the same principles as detailed above.

### Cross validation

Deep learning models were fit within IEL using stratified *k*-fold cross validation i.e. every one of the 100 models in each learning generation within IEL was individually trained and validated using cross-validation in the training partition. IEL allows the number of features used to fit each model to differ within each model in every generation. Accordingly, *k* (the number of splits) was set as the nearest integer above [sample size/number of features]. Cross validation was implemented with the scikit-learn StratifiedKFold function.

### Testing for generalization in holdout, unseen test data and performance measurement

After training was completed, optimized models generated by IEL were tested on the holdout, unseen test set for each sample and mental health condition by applying the requisite hyperparameter settings and selected features obtained from the 100 best-performing models in the training phase to the test set. The area under the receiver operating curve (AUROC), accuracy, precision, and recall were computed for test set models using standard Sci-Kit learn libraries and models with the best performance in each statistic selected for presentation as the final, optimized models. The threshold for prediction probability was 0.5 and receiver operating characteristic (ROC) curves are also provided for each experiment (**Supplementary Figures 1** and **2**).

### Feature importance determination

Shapley Additive Explanations (SHAP) values were computed using the SHAP toolbox (https://shap.readthedocs.io/en/latest/) to determine the relative importance of each feature to predicting cases of mental illness. SHAP is a game theoretic approach commonly used in ML to explain the output of any ML model including ‘black box’ estimators such as artificial neural networks and is considered resistant to multicollinearity. (45) It unifies prior methods such as LIME, Shapley sampling values and Tree Interpreter.

## RESULTS

### Overview

All results are from testing the final model obtained after optimization with IEL for generalization in the holdout, unseen test dataset for each participant sample and experiment. For each condition (depression, anxiety, SSD) a parallel set of results is presented for each participant sample of new onset cases at 11-12 yrs; all prevailing cases at 9-10 yrs and all prevailing cases at 11-12 yrs. In all experiments only data collected at 9-10 yrs is input to deep learning to make predictions. Thus, results obtained for new onset and prevailing cases at 11-12 yrs represent predictions of future case status.

For each disorder and age group, results are presented for the metrics below for a) <u>multimodal</u> models constructed using all types of input features; and b) <u>neural-only</u> models.

- Performance statistics: accuracy, precision, recall and AUROC. ROC curves may be viewed in **Supplementary Figures 1** and **2**.
- Final predictors ranked in order of importance by their group-level SHAP score (average absolute value across the participant sample) and the mean predictor importance for the requisite experiment.
- Individual-level final predictor importance (SHAP scores) across the participant sample. This summary plot is also used to determine the directionality of the relationship between the predictor and case status.

### Depression

Deep learning optimized with IEL predicted depression in early adolescence with >80% accuracy and recall and ≥90% AUROC across all experiments (**Table 5a**), with precision of ∼75-80%. Performance was slightly worse by a few percentage points in predicting new onset cases in the future (at 11-12 yrs) than either contemporaneous or all prevailing cases at 11-12 yrs. When each experiment was recapitulated using only neural candidate predictors, we found that final optimized predictive models displayed substantially lower performance (**Table 5b**) than those obtained with multimodal predictors with accuracy of ∼60-70% and AUROC of ∼60-77%, or some 10-25 percentage points lower than with multimodal predictors. Similar differentials were seen in precision and recall. In depression, multimodal models achieved somewhat better performance when predicting prevailing cases at 9-10yrs and11-12 vs new onset cases at 11-12 yrs. In neural-only models this was reversed, with a substantially stronger model obtained for new onset cases.

**Table 5:** Performance of deep learning optimized with Integrated Evolutionary Learning in predicting cases of depression using multimodal and neural-only feature types. Performance statistics of accuracy, precision, recall and the AUROC are shown for the most accurate model obtained with deep learning optimized with Integrated Evolutionary Learning using a) multimodal features and b) only neural features. We used features obtained at 9-10 years of age to predict new onset cases of depression at 11-12 years of age as well as all prevailing contemporaneous cases (9-10 yrs) and all prevailing cases at 11-12 years of age. Corresponding ROC curves may be viewed in **Supplementary Figures 1** and **2**.

| Age of case determination | Accuracy (%) | Precision (%) | Recall (%) | AUC |
| --- | --- | --- | --- | --- |
| New onset at age 11-12 years | 81.3 | 75.0 | 84.0 | 89.5 |
| All cases at age 9-10 years | 85.3 | 78.4 | 87.3 | 92.0 |
| All cases at age 11-12 years | 85.7 | 79.8 | 89.3 | 91.1 |

**Table 5: Performance of deep learning optimized with Integrated Evolutionary Learning in predicting cases of depression using multimodal and neural-only feature types**
| Age of case determination | Accuracy (%) | Precision (%) | Recall (%) | AUC |
| --- | --- | --- | --- | --- |
| New onset at age 11-12 years | 69.6 | 63.5 | 71.4 | 76.8 |
| All cases at age 9-10 years | 61.3 | 56.8 | 66.7 | 60.6 |
| All cases at age 11-12 years | 61.9 | 58.3 | 41.7 | 59.5 |

In interpreting multimodal models (**Table 6**), we found that parent problem behaviors were the most important predictor of early adolescent depression in each participant sample. Specific parental behavioral drivers of youth cases differed by age and case type. In new onset cases at 11-12 yrs, parent externalizing traits were the most important predictors vs the total burden of parental behavioral health problems in all prevailing cases at 11-12 yrs. In contemporaneous cases at 9-10 yrs, parent avoidant and intrusive traits appeared as final predictors. Sleep disturbances meeting clinical criteria were important in predicting prevailing cases of depression at 9-10 and 11-12 yrs, but not in predicting new onset cases. In the latter, acceptance by a secondary caregiver and prosocial behaviors had an inverse relationship with depression onset. Interestingly, the second most important predictor of new onset cases was whether the child had ever previously received mental health or substance abuse services, suggesting these children had already come to clinical attention at or before 9-10 yrs and the onset of depression. Group-level importances for multimodal model predictors (averaged across the participant sample) were in the range [0.14, 0.21] and the mean importance for each experiment in the range [0.10, 0.16].

**Table 6:** Final predictors of cases of depression in early adolescence. Final predictors of cases of all prevailing cases of depression at ages 9-10 and 11-12 years as well as new onset cases only at 11-12 years of age are shown for the most accurate models obtained using deep learning optimized with IEL obtained with a) multimodal features and b) only neural features. Final predictors are ranked in order of importance where the relative importance of each predictor is computed with the Shapley Additive Explanations technique and presented here averaged across all participants in the sample. Features in blue indicate an inverse relationship with depression verified with the Shapley method. MH = mental health; SU=substance use; SST = Standard Stop Signal task; MID = Monetary Incentive Delay task; ROI = region of interest; FA = fractional anisotropy; LD = longitudinal diffusivity; WM = white matter; GM = gray matter.

| Age of case determination | Ranked Final Predictors | Importance |
| --- | --- | --- |
| New onset at age 11-12 years | Parent externalizing scale score | 0.20 |
|  | Ever received MH/SU services | 0.12 |
|  | Secondary caregiver acceptance | 0.05 |
|  | Prosocial behaviors | 0.04 |
|  | Mean | 0.10 |
| All cases at age 9-10 years | Parent avoidant personality problems | 0.21 |
|  | Sleep disorder of excessive somnolence | 0.16 |
|  | Parent intrusive syndrome score | 0.05 |
|  | Mean | 0.14 |
| All cases at age 11-12 years | Parent total problems syndrome score | 0.24 |
|  | Total sleep disturbances scale score | 0.08 |
|  | Mean | 0.16 |

**Table 6: Final predictors of cases of depression in early adolescence**
| Neural data type | Ranked Final Predictors | Importance |
| --- | --- | --- |
| New onset at age 11-12 years | SST incorrect vs correct go contrast left caudal anterior cingulate ROI | 0.018 |
|  | SST any stop vs correct go contrast left pars opercularis ROI | 0.017 |
|  | T1 WM intensity for genetic parcel right central hemisphere | 0.014 |
|  | SST incorrect stop vs correct go contrast left transverse temporal ROI | 0.013 |
|  | T1 GM intensity right precentral ROI | 0.012 |
|  | FA in right lateral orbital frontal GM ROI | 0.009 |
|  | SST correct go vs fixation contrast in left entorhinal ROI | 0.008 |
|  | SST incorrect go vs correct go contrast in left accumbens ROI | 0.008 |
|  | SST incorrect stop vs correct go contrast in right parstriangularis ROI | 0.006 |
|  | Mean | 0.012 |
| All cases at age 9-10 years | FA in GM in right rostral middle frontal ROI | 0.011 |
|  | LD in GM in left banks of superior temporal sulcus ROI | 0.010 |
|  | Correlation between dorsal attention and default mode networks | 0.007 |
|  | SST correct vs incorrect stop contrast in right transversetemporal ROI | 0.005 |
|  | SST average framewise rotation in radians | 0.004 |
|  | Correlation between default mode and sensorimotor hand networks | 0.003 |
|  | SST maximum framewise rotation in radians | 0.003 |
|  | Mean | 0.006 |
| All cases at age 11-12 years | MID anticipation of small loss vs neutral contrast in right inferior temporal ROI | 0.028 |
|  | SST average framewise rotation in radians | 0.006 |
|  | Mean | 0.017 |

Final predictors of new onset cases at 11-12 yrs obtained in neural-only models (**Table 6b**) were dominated by features derived from the Standard Stop Signal fMRI task, which measures response inhibition. Here, SST ROIs emphasized the left hemisphere. Specifically, SST responses in pars opercularis and (right) pars triangularis (collectively, Broca’s area), caudal anterior cingulate and entorhinal ROIs exhibited inverse directionality with depression where ROIs in the transverse temporal (Heschl’s gyri) and accumbens were positively related to depression onset. Certain structural metrics also appeared as final predictors of new onset cases. Specifically, right precentral and lateral orbital frontal gray matter ROIs and white matter intensity in a genetically-defined right hemisphere parcel.

Gray matter structural features in right rostral middle frontal and left superior temporal sulcus ROIs were prominent in predicting contemporaneous prevailing cases of depression at 9-10yrs as were correlation strengths between the dorsal attention and default mode networks and default mode and sensorimotor hand networks. While SST contrast in the right transverse temporal ROI had an inverse relationship with cases status at this age, we note that head motion during the SST task also appeared. The final predictive model for all prevailing future cases of depression at 11-12 yrs was parsimonious and only comprised contrast differences in the right inferior temporal ROI in the Monetary Incentive Delay task, which measures approach and avoidance during reward processing, and a metric of head motion (framewise displacement) in the SST task. Group-level importances for neural-only model predictors were in the range [0.003,0.028] and the mean importance for each experiment in the range [0.006,0.017], both representing lower importance ranges than multimodal models.

Where **Table 6** presents the importance of final predictors as summarized (mean absolute value) across the requisite experimental participant sample, we were also interested in predictor importance at the individual participant level. We computed and plotted individual-level SHAP values to understand both the dispersion of predictor importances across individuals and the directionality of the relationship between final predictors and clinical case status (**Figure 3**). In SHAP summary plots, each data point represents an individual participant and the colorization reflects the original value of the predictor as an input feature. Thus, discrete-valued features appear as red or blue, whereas a continuous feature appears as a color gradient from low to high. The directionality of the relationship between predictors and depression case status obtained in these plots was further compared with coefficients obtained during LASSO regression for **Coarse Feature Selection** (**Supplementary Table 3**) and found to be in agreement.

**Figure 3:**
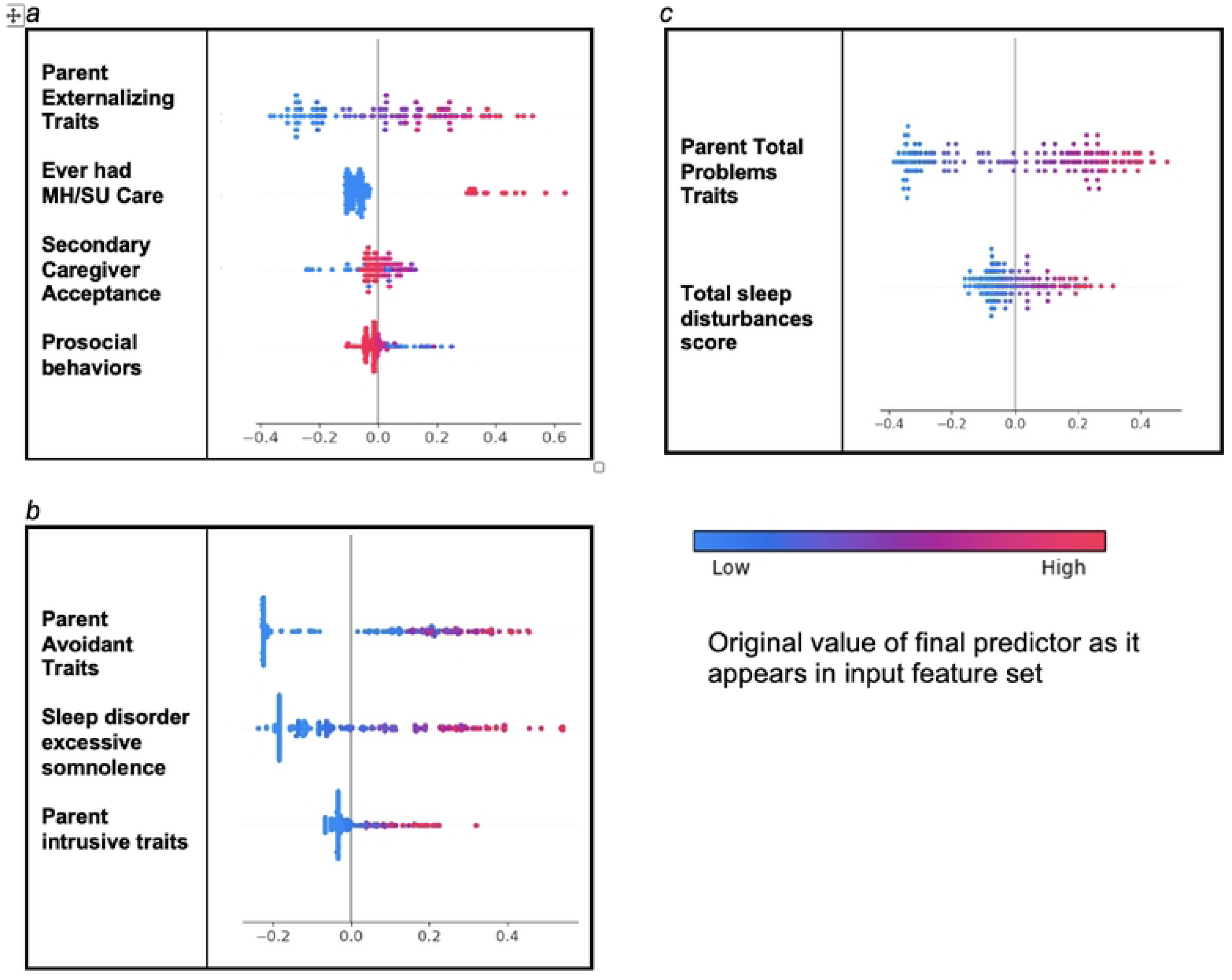
Individual-level importances of depression predictors in multimodal predictive models. Summary plots are presented of the importance of each final predictor (computed with the Shapley Additive Explanations technique) on an individual subject level to predicting depression a) with new onset at 11-12 yrs; b) in all cases at 9-10 yrs; and c) in all cases at 11-12 yrs. The color gradient represents the original value of each feature (metric) where red = high and blue = low. Discrete (binary) features appear as red or blue, while continuous features appear as a color gradient from low to high.

**Figure 3** reveals that individual-level importance of final predictors in early adolescent depression are typically widely dispersed. For example, when predicting new onset cases of depression at 11-12 yrs, the leading predictor of parent externalizing traits has a large range of ∼[-0.4,0.6] across individual participants. Further, dispersion is typically greater for the more important predictors. Overall, these plots also indicate that all final predictors obtained have a positive relationship with depression case status, with the exception of secondary caregiver acceptance and prosocial behaviors in predicting new onset cases (see also **Table 6**). We also computed individual-level importances of final predictors for neural-only experiments (**Figure 4**). Here, the dispersion of individual-level predictor importances across participants were consistently smaller in neural-only versus multimodal prediction of early adolescent depression.

**Figure 4:**
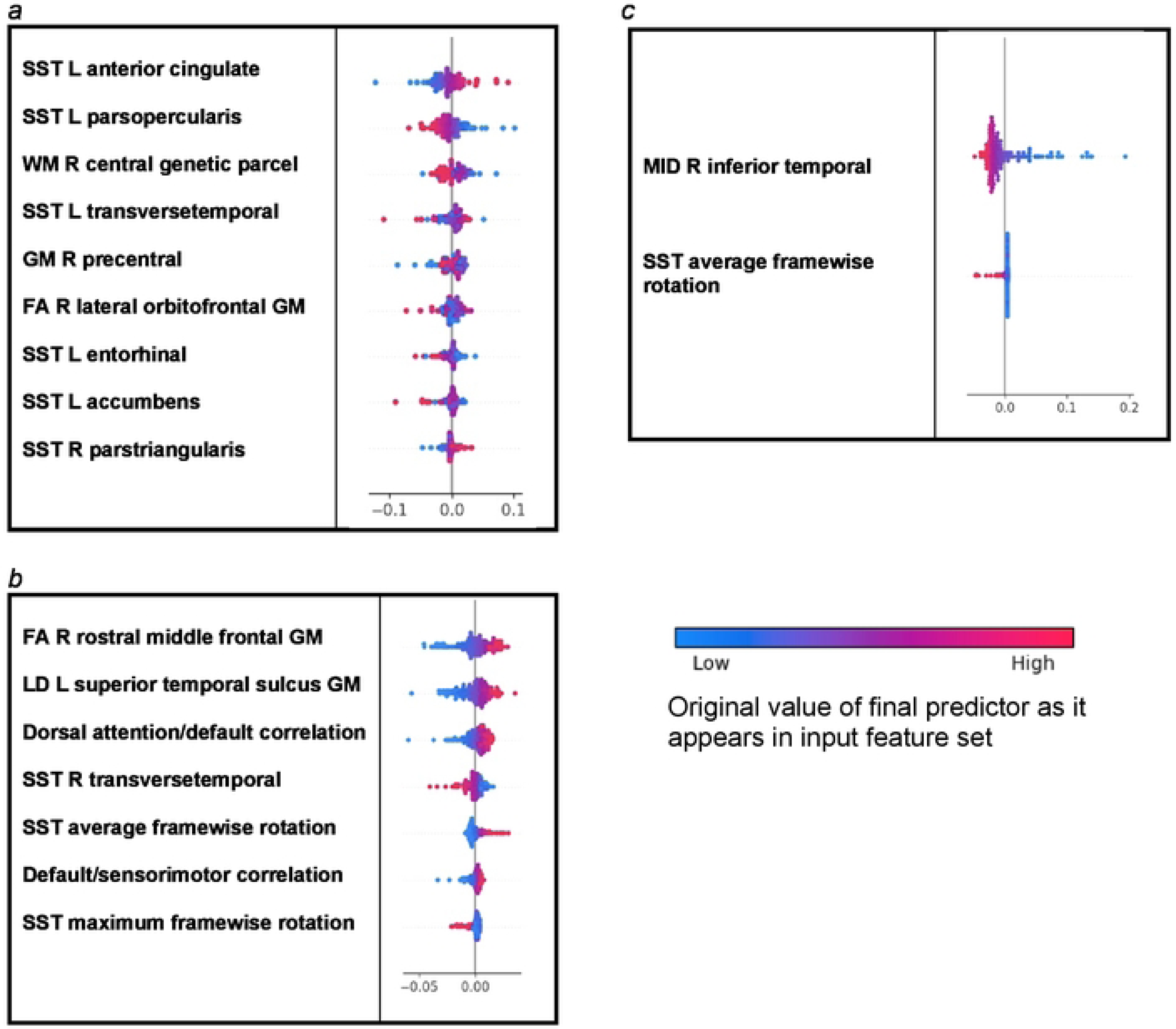
Individual-level importances of depression predictors in neural-only predictive models. Summary plots are presented of the importance of each final predictor (computed with the Shapley Additive Explanations technique) on an individual subject level to predicting depression a) with new onset at 11-12 yrs; in all cases at 9-10 yrs; and c) in all cases at 11-12 yrs. The color gradient represents the original value of each feature (metric) where red = high and blue = low. Discrete (binary) features appear as red or blue, while continuous features appear as a color gradient.

### Anxiety

Deep learning optimized with IEL performed very well in predicting both new onset and prevailing cases of anxiety in early adolescence. In anxiety, ∼93% accuracy and ∼96% AUROC was achieved in predicting new onset cases versus ∼85% accuracy and ∼91% AUROC in predicting prevailing cases using data obtained at 9-10 yrs to predict cases at the future time point of 11-12 yrs. The best overall performance was observed using data at 9-10 yrs to predict contemporaneous prevailing cases, with ∼97% accuracy and nearly 100% AUROC achieved (**Table 7a**). Similar to depression, neural-only models did not perform as well as multimodal models in predicting anxiety cases, being ∼20-40% less accurate. Best performance was obtained when predicting new onset anxiety at 11-12 yrs, where the final, optimized neural-only model achieved 75% accuracy and ∼78% AUROC. In comparison, neural-only predictive models of all prevailing cases at 9-10 yrs and 11-12 yrs showed substantially inferior performance with accuracy of ∼58 and ∼64% and AUROC of 57 and 63% respectively (**Table 7b**)

**Table 7:** Performance of deep learning optimized with IEL in predicting cases of anxiety. Performance statistics of accuracy, precision, recall and the AUROC are shown for the most accurate model obtained with deep learning optimized with Integrated Evolutionary Learning using a) multimodal features and b) only neural features. We used features obtained at 9-10 years of age to predict new onset cases of anxiety at 11-12 years of age as well as all prevailing contemporaneous cases (9-10 yrs) and all prevailing cases at 11-12 years of age. Corresponding ROC curves may be viewed in **Supplementary Figures 1** and **2**.

| Age of case determination | Accuracy (%) | Precision (%) | Recall (%) | AUC |
| --- | --- | --- | --- | --- |
| New onset at age 11-12 years | 93.3 | 93.3 | 86.7 | 95.9 |
| All cases at age 9-10 years | 96.5 | 95.2 | 95.6 | 99.5 |
| All cases at age 11-12 years | 84.8 | 80.2 | 82.1 | 90.5 |

**Table 7: Performance of deep learning optimized with IEL in predicting cases of anxiety**
| Neural data type | Accuracy (%) | Precision (%) | Recall (%) | AUC |
| --- | --- | --- | --- | --- |
| New onset at age 11-12 years | 75.0 | 70.7 | 63.3 | 77.6 |
| All cases at age 9-10 years | 57.9 | 54.4 | 72.8 | 57.0 |
| All cases at age 11-12 years | 63.8 | 59.2 | 55.4 | 63.0 |

In anxiety, new onset cases were predicted with a relatively complex final model comprising 8 predictors (**Table 8a**). Here, the most important predictor was whether the youth had previously come to clinical attention (ever received mental health or substance use services), closely followed by the youth’s total burden of clinically-significant sleep disturbances and whether the child’s mother had received clinical treatment for a mental or emotional problem. The degree of parent externalizing and avoidant behavioral problems was also important. These were followed by three less important predictors with an inverse relationship with case status: loss contrast in the left orbitofrontal ROI in the Monetary Incentive Delay task, whether parent and youth got along very well and the youth’s prosocial scale score. The appearance of MID contrast in the left orbitofrontal (OFC) ROI is of particular note since this was the only model in which a neural feature survived the large-scale, parallelized optimization process to appear as a final predictor in a multimodal analysis.

**Table 8:** Final predictors of cases of anxiety in early adolescence. Final predictors of cases of all prevailing cases of anxiety at ages 9-10 and 11-12 years as well as new onset cases only at 11-12 years of age are shown for the most accurate models obtained using deep learning optimized with IEL obtained with a) multimodal features and b) only neural features. Final predictors are ranked in order of importance where the relative importance of each predictor is computed with the Shapley Additive Explanations technique and presented here averaged across all participants in the sample. Features in blue indicate an inverse relationship with depression verified with the Shapley method. MH = mental health; SU=substance use; SST = Standard Stop Signal task; MID = Monetary Incentive Delay task; ROI = region of interest; FA = fractional anisotropy; LD = longitudinal diffusivity; WM = white matter; GM = gray matter.

| Age of case determination | Ranked Final Predictors | Importance |
| --- | --- | --- |
| New onset at age 11-12 years | Ever received MH/SU services | 0.12 |
|  | Total sleep disturbance | 0.11 |
|  | Mother been to a counselor due to mental or emotional problem | 0.10 |
|  | Parent externalizing syndrome score | 0.08 |
|  | Parent avoidant syndrome score | 0.06 |
|  | MID large vs small loss contrast in left orbitofrontal ROI | 0.05 |
|  | Parent and youth get along very well | 0.05 |
|  | Prosocial behaviors scale score | 0.03 |
|  | **Mean** | 0.08 |
| All cases at age 9-10 years | Total sleep disturbances scale score | 0.17 |
|  | Parent total problems syndrome score | 0.10 |
|  | Ever received MH/SU services | 0.08 |
|  | Parent anxiety syndrome score | 0.06 |
|  | Sleep disorders of arousal | 0.05 |
|  | Parent and youth get along very well | 0.04 |
|  | Has more than 3 friends in regular friend group | 0.02 |
|  | Disorders of initiating and maintaining sleep | 0.02 |
|  | **Mean** | 0.07 |
| All cases at age 11-12 years | Parent total problems syndrome score | 0.23 |
|  | Mother been to a counselor due to mental or emotional problem | 0.09 |
|  | Parent and youth get along very well | 0.05 |
|  | **Mean** | 0.12 |

**Table 8: Final predictors of cases of anxiety in early adolescence**
| Neural data type | Ranked Final Predictors | Importance |
| --- | --- | --- |
| **New onset at age 11-12 years** | MID large vs small loss contrast left medial orbitofrontal ROI<br>MID large vs small loss contrast left ventral diencephalon<br>T1 intensity right inferior lateral ventricle ROI<br>T1 white-gray contrast left precuneus ROI<br>T1 white-gray contrast right paracentral ROI<br>Mean cortical sulcal depth in mm for left hemisphere<br>**Mean** | 0.110<br>0.075<br>0.070<br>0.056<br>0.051<br>0.028<br>0.065 |
| **All cases at age 9-10 years** | FA in sub-adjacent WM associated with cortical right temporal pole ROI<br>FA in GM associated with left fusiform ROI<br>nBack 2 back condition in left transverse temporal ROI<br>**Mean** | 0.004<br>0.003<br>0.002<br>0.003 |
| **All cases at age 11-12 years** | Weighted average for genetic parcellation in right orbitofrontal<br>FA in sub-adjacent WM associated with right entorhinal ROI<br>MID anticipation of small loss vs neutral contrast in left lateral ventricle ROI<br>T1 white-gray contrast in right frontal pole ROI<br>Cortical thickness in mm of right caudal anterior cingulate ROI<br>MID all loss positive vs negative feedback contrast in left inferior temporal ROI<br>**Mean** | 0.035<br>0.034<br>0.027<br>0.021<br>0.007<br>0.004<br>0.021 |

We detected overlap between the final predictors of new onset cases of anxiety at 11-12 yrs and those which predicted prevailing cases at 9-10 and 11-12 yrs. Sleep disturbance (total and disorders of initiating and maintaining sleep) was similarly prominent in predicting contemporaneous cases but here parent behavioral factors isolated as final predictors were anxiety traits and the parent total burden of behavioral problems. Predictors exhibiting an inverse relationship with case status at 9-10 yrs were how well parent and youth got along (as with new onset cases) and whether the youth had >3 friends in their regular group. The model predicting all prevailing cases of anxiety at 11-12 yrs was more parsimonious, with three final predictors of parent total behavioral problems, the mother’s history of clinical treatment and whether parent and child got along well. Group-level importances for multimodal model predictors were in the range [0.02, 0.23] and the mean importance for each experiment in the range [0.07, 0.12].

In neural-only models predicting new onset anxiety cases, features from the MID fMRI task and structural metrics predominated (**Table 8b**). The most important final predictors were MID contrast in the left medial OFC ROI and ventral diencephalon and T1 intensity in the right inferior lateral ventricle (temporal horn). All had an inverse relationship with case status. Further final predictors with a positive relationship with anxiety were all structural: white-gray matter contrast in the left precuneus ROI and right paracentral ROI and mean cortical sulcal depth in mm for the left hemisphere as a whole. As noted above, neural-only predictive models of prevailing anxiety cases at 9-10 yrs and 11-12 yrs were substantially less reliable with smaller mean predictor importances vs new onset cases. Final, optimized models predicting prevailing cases at 11-12 yrs emphasized features from the MID task and structural metrics. Group-level importances for neural-only model predictors were in the range [0.002, 0.11] and the mean importance for each experiment in the range [0.003, 0.065].

To probe the dispersion of predictor importances at the individual level, we again developed summary plots of individual-level importances (**Figures 5** and **6)**. Similarly to depression, we observed relatively more widely dispersed individual-level importances over the participant sample in multimodal vs neural-only models, and the trend for wider dispersion of predictor importance in the more important final predictors. The directionality of the relationship between predictors and depression case status obtained in these plots was further compared with coefficients obtained during LASSO regression for **Coarse Feature Selection** (**Supplementary Table 3**) and found to be in agreement.

**Figure 5:**
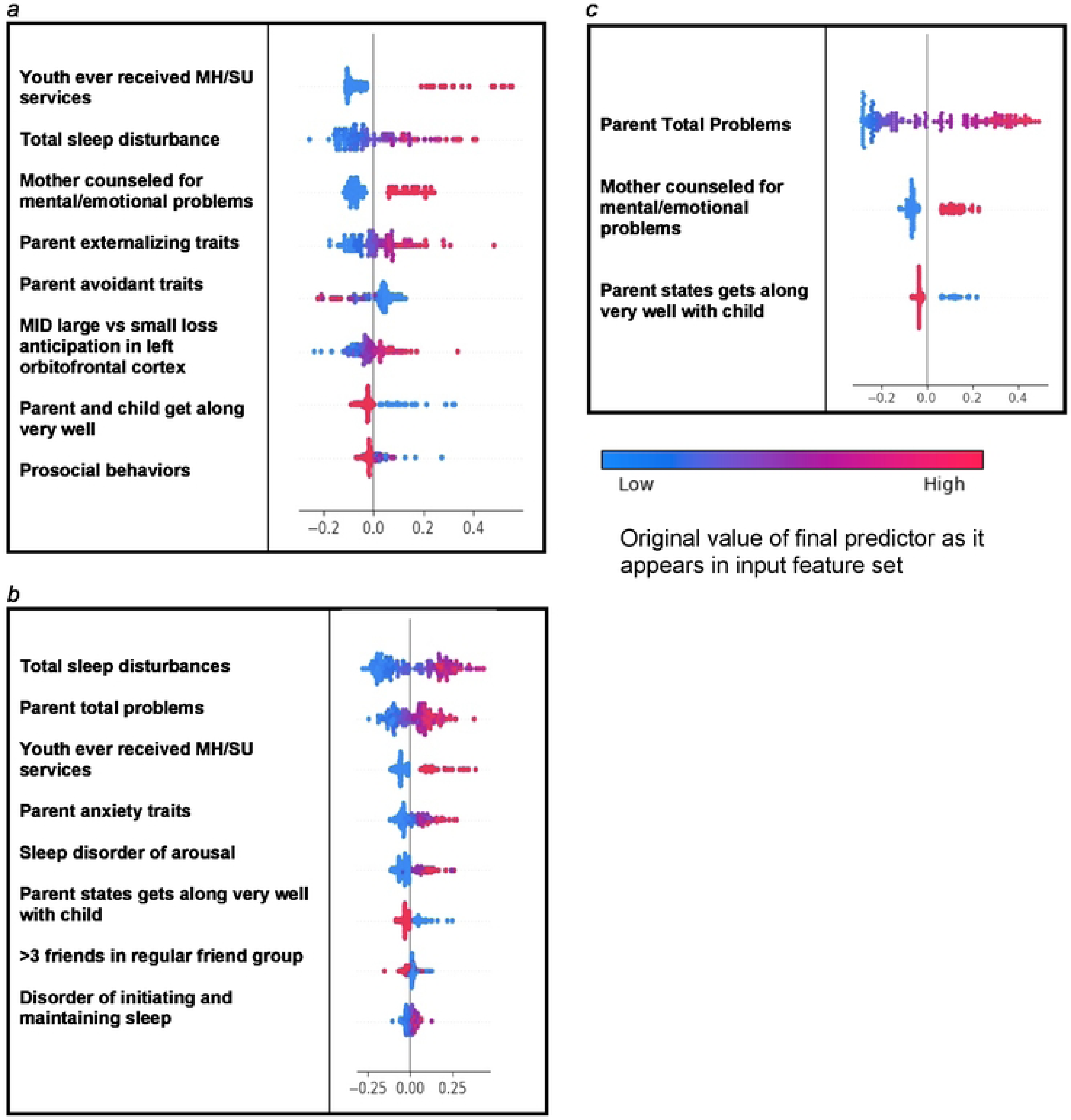
Individual-level importances of final predictors of anxiety in early adolescence. Summary plots are presented of the importance of each final predictor (computed with the Shapley Additive Explanations technique) on an individual subject level to predicting anxiety a) with new onset at 11-12 yrs; b) in all cases at 9-10 yrs; and c) in all cases at 11-12 yrs. The color gradient represents the original value of each feature (metric) where red = high and blue = low. Discrete (binary) features appear as red or blue, while continuous features appear as a color gradient.

**Figure 6:**
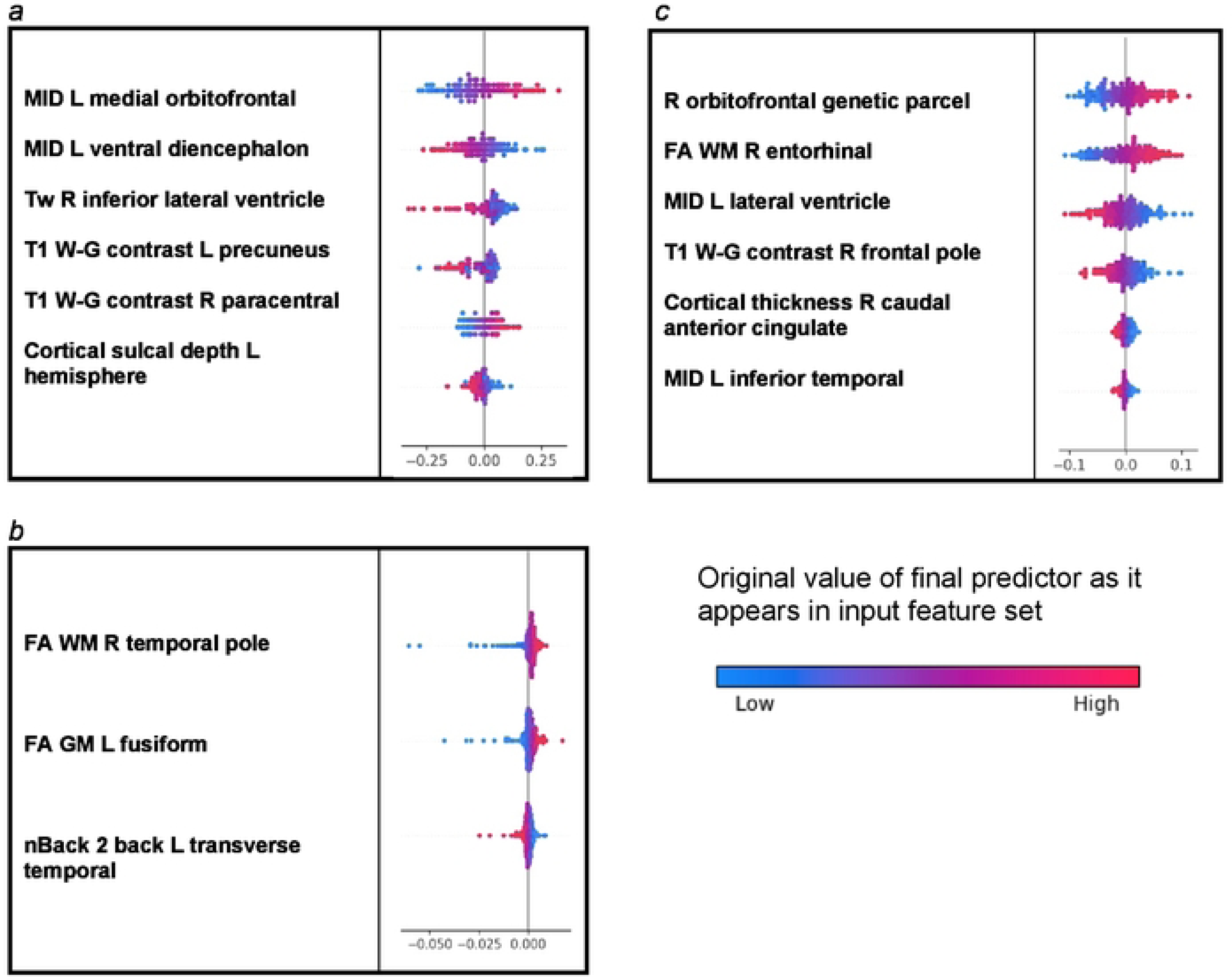
Individual-level importances of neural final predictors of anxiety in early adolescence. Summary plots are presented of the importance of each final predictor (computed with the Shapley Additive Explanations technique) on an individual subject level to predicting anxiety a) with new onset at 11-12 yrs; b) in all cases at 9-10 yrs; and c) in all cases at 11-12 yrs. The color gradient represents the original value of each feature (metric) where red = high and blue = low. Discrete (binary) features appear as red or blue, while continuous features appear as a color gradient.

Individual-level predictor importances for the best-performing mixed-type neural models of anxiety again showed reduced dispersion across the participant group (**Figure 6**) when compared with multimodal models (**Figure 5**). The widest dispersion was observed when predicting new onset cases of anxiety.

### Somatic Symptom Disorder

Deep learning optimized with IEL performed well using multimodal data in predicting both new onset and prevailing cases of SSD in early adolescence. Here, ∼84% accuracy and ∼89% AUROC was achieved in predicting future, new onset cases at 11-12 yrs with data obtained at 9-10 yrs. The best overall performance was observed using data at 9-10 yrs to predict contemporaneous prevailing cases, with ∼95% accuracy and ∼98% AUROC. Predictive performance of all prevailing cases at 11-12 yrs using data from 9-10yrs was comparable to new onset predictions, with accuracy of ∼84% and AUROC of ∼92% (**Table 9a**). As with depression and anxiety, neural-only models did not perform as well as multimodal models (**Table 9b**), being ∼10-25% less accurate. The best performance was seen in predicting new onset cases at 11-12 yrs with accuracy of ∼67% and AUROC of ∼66% and all prevailing cases at 9-10 yrs with accuracy of ∼67% and AUROC of ∼72%. Accuracy in the model predicting prevailing cases at 11-12 yrs dropped to ∼63% with a similar AUROC.

**Table 9:** Performance of deep learning optimized with Integrated Evolutionary Learning in predicting cases of Somatic Symptom Disorder. Performance statistics of accuracy, precision, recall and the AUC are shown for the most accurate model obtained with deep learning optimized with Integrated Evolutionary Learning using a) multimodal features and b) only neural features. We used features obtained at 9-10 years of age to predict new onset cases of somatic symptom disorder at 11-12 years of age as well as all prevailing contemporaneous cases (9-10 yrs) and all prevailing cases at 11-12 years of age. Corresponding ROC curves may be viewed in **Supplementary Figures 1** and **2**.

| Age of case determination | Accuracy (%) | Precision (%) | Recall (%) | AUC |
| --- | --- | --- | --- | --- |
| New onset at age 11-12 years | 83.6 | 78.8 | 80.6 | 88.7 |
| All cases at age 9-10 years | 94.5 | 93.8 | 90.6 | 98.4 |
| All cases at age 11-12 years | 83.5 | 77.2 | 87.1 | 91.5 |

**Table 9: Performance of deep learning optimized with Integrated Evolutionary Learning in predicting cases of Somatic Symptom Disorder**
| Neural data type | Accuracy (%) | Precision (%) | Recall (%) | AUC |
| --- | --- | --- | --- | --- |
| New onset at age 11-12 years | 67.2 | 65.0 | 40.3 | 65.8 |
| All cases at age 9-10 years | 67.2 | 63.1 | 50.0 | 71.7 |
| All cases at age 11-12 years | 62.5 | 58.9 | 41.9 | 63.7 |

In interpreting optimized multimodal predictive models for early adolescent SSD we observed that new onset cases were predicted by the level of total sleep disturbance, parent somatization score on the adult CBCL and whether the child had ever received mental health clinical services (**Table 10a**). While sets of specific predictors were not the same, overlap was observed among age groups. Of note, parental level of somatization predicted both new onset cases and contemporaneous prevailing cases at 9-10 yrs. In addition, sleep disturbances of various types were a common theme across all three age groups. The highly accurate model predicting cases at 9-10 yrs was also interesting in featuring whether the child had seen a clinician for a medical issue other than a regular checkup and whether parent and child got along very well. The latter was the only final predictor with an inverse relationship with SSD. Group-level importances for multimodal model predictors were in the range [0.02, 0.27] and the mean importance for each experiment in the range [0.09, 0.16].

**Table 10:**
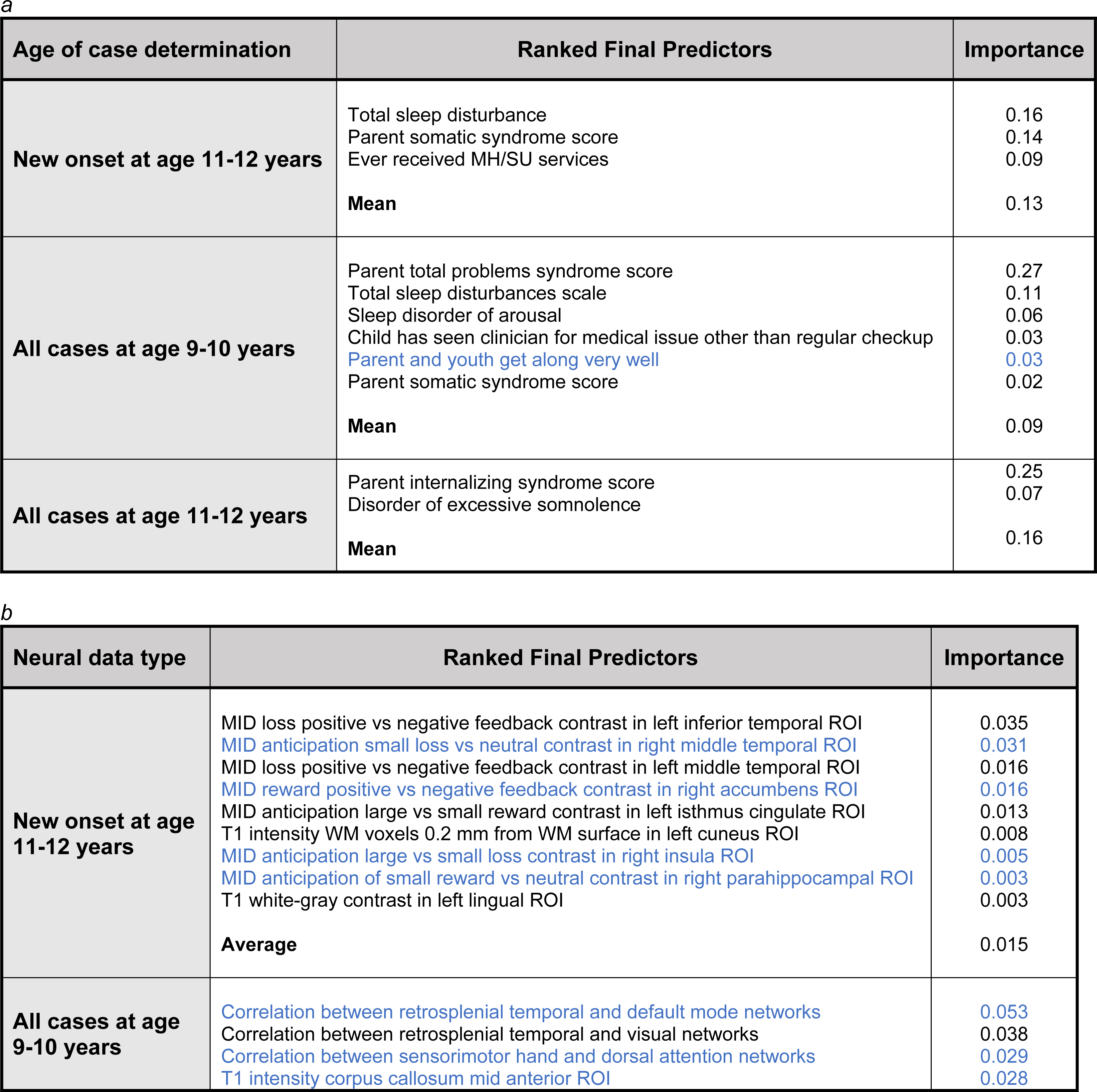
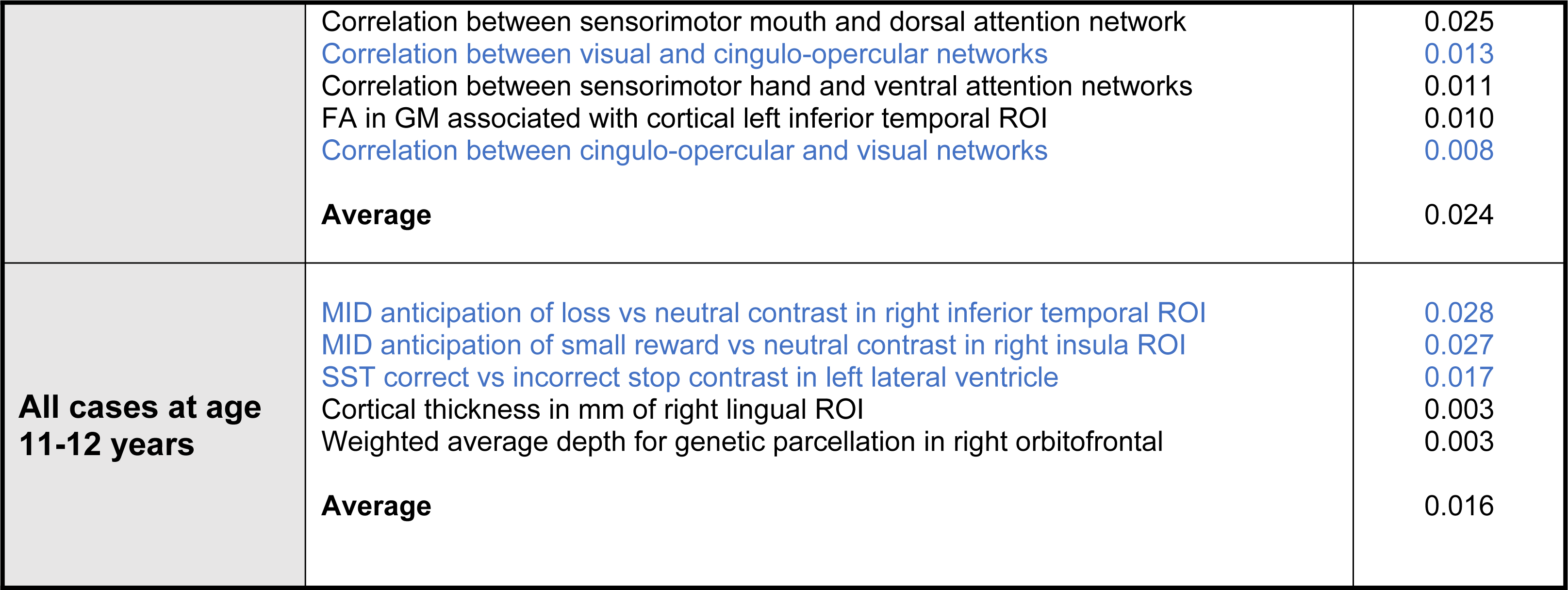
Final predictors of cases of somatic symptom disorder in early adolescence. Final predictors of cases of all prevailing cases of SSD at ages 9-10 and 11-12 years as well as new onset cases only at 11-12 years of age are shown for the most accurate models obtained using deep learning optimized with IEL obtained with a) multimodal features and b) only neural features. Final predictors are ranked in order of importance where the relative importance of each predictor is computed with the Shapley Additive Explanations technique and presented here averaged across all participants in the sample. Features in blue indicate an inverse relationship with depression verified with the Shapley method. MH = mental health; SU=substance use; SST = Standard Stop Signal task; MID = Monetary Incentive Delay task; ROI = region of interest; FA = fractional anisotropy; LD = longitudinal diffusivity; WM = white matter; GM = gray matter.

| Age of case determination | Ranked Final Predictors | Importance |
| --- | --- | --- |
| New onset at age 11-12 years | Total sleep disturbance | 0.16 |
|  | Parent somatic syndrome score | 0.14 |
|  | Ever received MH/SU services | 0.09 |
|  | **Mean** | 0.13 |
| All cases at age 9-10 years | Parent total problems syndrome score | 0.27 |
|  | Total sleep disturbances scale | 0.11 |
|  | Sleep disorder of arousal | 0.06 |
|  | Child has seen clinician for medical issue other than regular checkup | 0.03 |
|  | Parent and youth get along very well | 0.03 |
|  | Parent somatic syndrome score | 0.02 |
|  | **Mean** | 0.09 |
| All cases at age 11-12 years | Parent internalizing syndrome score | 0.25 |
|  | Disorder of excessive somnolence | 0.07 |
|  | **Mean** | 0.16 |

**Table 10: Final predictors of cases of somatic symptom disorder in early adolescence**
| Neural data type | Ranked Final Predictors | Importance |
| --- | --- | --- |
| New onset at age 11-12 years | MID loss positive vs negative feedback contrast in left inferior temporal ROI | 0.035 |
|  | MID anticipation small loss vs neutral contrast in right middle temporal ROI | 0.031 |
|  | MID loss positive vs negative feedback contrast in left middle temporal ROI | 0.016 |
|  | MID reward positive vs negative feedback contrast in right accumbens ROI | 0.016 |
|  | MID anticipation large vs small reward contrast in left isthmus cingulate ROI | 0.013 |
|  | T1 intensity WM voxels 0.2 mm from WM surface in left cuneus ROI | 0.008 |
|  | MID anticipation large vs small loss contrast in right insula ROI | 0.005 |
|  | MID anticipation of small reward vs neutral contrast in right parahippocampal ROI | 0.003 |
|  | T1 white-gray contrast in left lingual ROI | 0.003 |
|  | **Average** | 0.015 |
| All cases at age 9-10 years | Correlation between retrosplenial temporal and default mode networks | 0.053 |
|  | Correlation between retrosplenial temporal and visual networks | 0.038 |
|  | Correlation between sensorimotor hand and dorsal attention networks | 0.029 |
|  | T1 intensity corpus callosum mid anterior ROI | 0.028 |
|  | Correlation between sensorimotor mouth and dorsal attention network | 0.025 |
|  | Correlation between visual and cingulo-opercular networks | 0.013 |
|  | Correlation between sensorimotor hand and ventral attention networks | 0.011 |
|  | FA in GM associated with cortical left inferior temporal ROI | 0.010 |
|  | Correlation between cingulo-opercular and visual networks | 0.008 |
|  | **Average** | 0.024 |
| **All cases at age 11-12 years** | MID anticipation of loss vs neutral contrast in right inferior temporal ROI | 0.028 |
|  | MID anticipation of small reward vs neutral contrast in right insula ROI | 0.027 |
|  | SST correct vs incorrect stop contrast in left lateral ventricle | 0.017 |
|  | Cortical thickness in mm of right lingual ROI | 0.003 |
|  | Weighted average depth for genetic parcellation in right orbitofrontal | 0.003 |
|  | **Average** | 0.016 |

In neural-only models, we found that MID fMRI task features were emphasized in predicting new onset cases. Interestingly, all MID features from the left hemisphere (inferior and middle temporal, isthmus cingulate ROIs) had a positive relationship with case status while those from the right hemisphere (middle temporal, accumbens, insula and parahippocampal ROIs) showed an inverse relationship with SSD (**Table 10b**, Figure 8). Other structural predictors of new onset cases were white matter intensity in the left cuneus ROI and white-gray contrast in the left lingual ROI. As with new onset cases, final predictors of prevailing cases of SSD at 11-12 yrs centered on the MID and structural neural features. Specific neural predictors of all prevailing cases at 11-12 yrs showed some commonality with new onset cases, with inverse relationships between MID contrast in the right inferior temporal and insula ROIs and case status. In contrast, the final, optimized model predicting all prevailing cases at 9-10 yrs was dominated by connectivity metrics derived from rsfMRI, evenly split between connectivity features with positive and inverse relationships with case status (Figure 8).

When examined at the individual level, final predictors of SSD in each participant sample showed the same patterns as we observed in depression and anxiety. Individual-level predictor importances were widely dispersed, where typically the more important predictors exhibited wider dispersions (Figures 7 and **8**). Further, the dispersion of individual-level importances was greater in the more accurate multimodal models.

**Figure 7:**
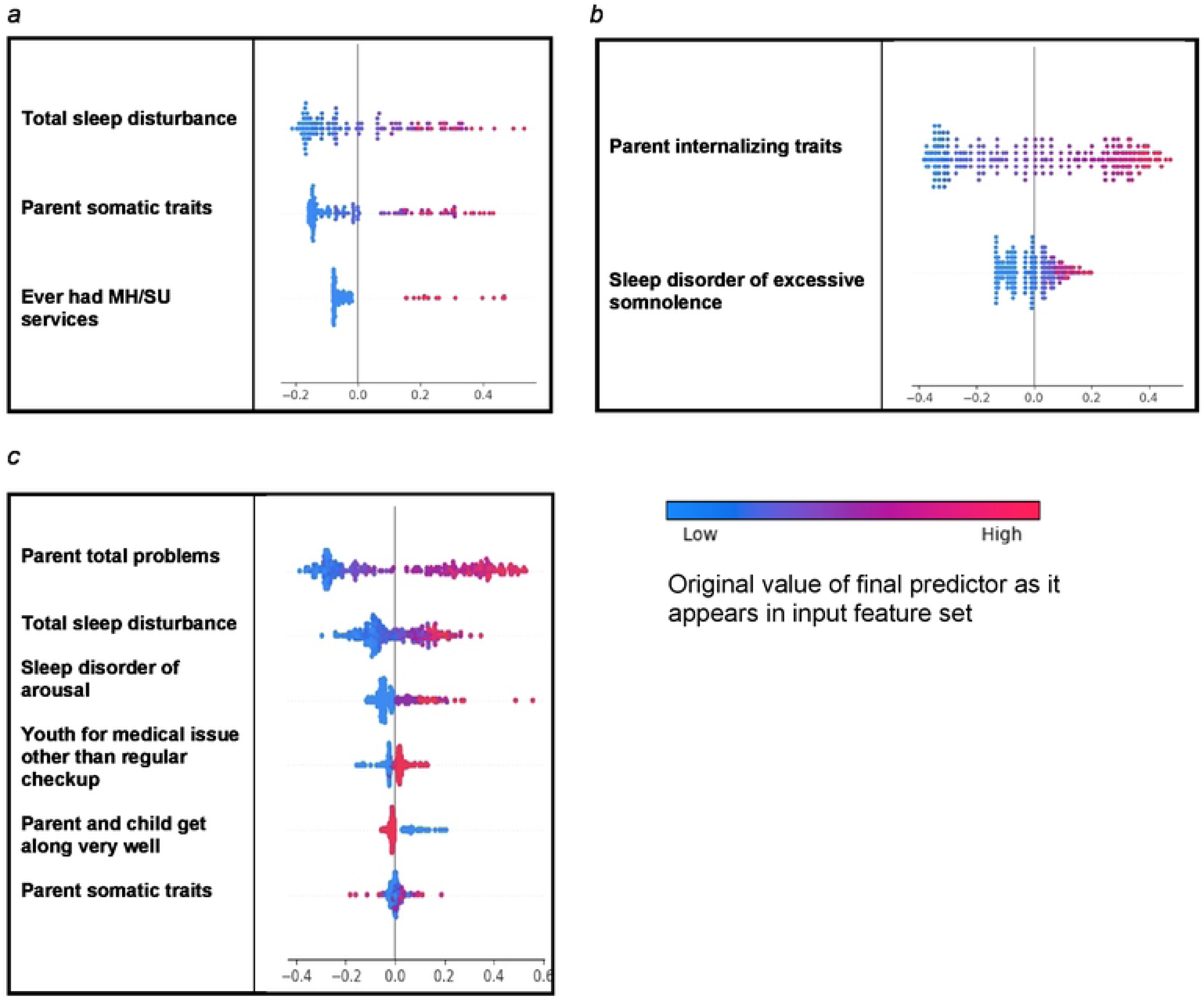
Individual-level importances of final predictors of somatic disorder in early adolescence. Summary plots are presented of the importance of each final predictor (computed with the Shapley Additive Explanations technique) on an individual subject level to predicting SSD a) with new onset at 11-12 yrs; b) in all cases at 9-10 yrs; and in all cases at 11-12 yrs. The color gradient represents the original value of each feature (metric) where red = high and blue = low. Discrete (binary) features appear as red or blue, while continuous features appear as a color gradient.

Similarly to depression and anxiety, individual-level importances of final predictors of somatic symptom disorder were less widely dispersed than multimodal models, being in the range [0.001, 0.025] and the more important predictors were more widely dispersed (Figure 8). The directionality of the relationship between predictors and SSD case status obtained in these plots was further compared with coefficients obtained during LASSO regression for **Coarse Feature Selection** (**Supplementary Table 3**) and found to be in agreement.

**Figure 8:**
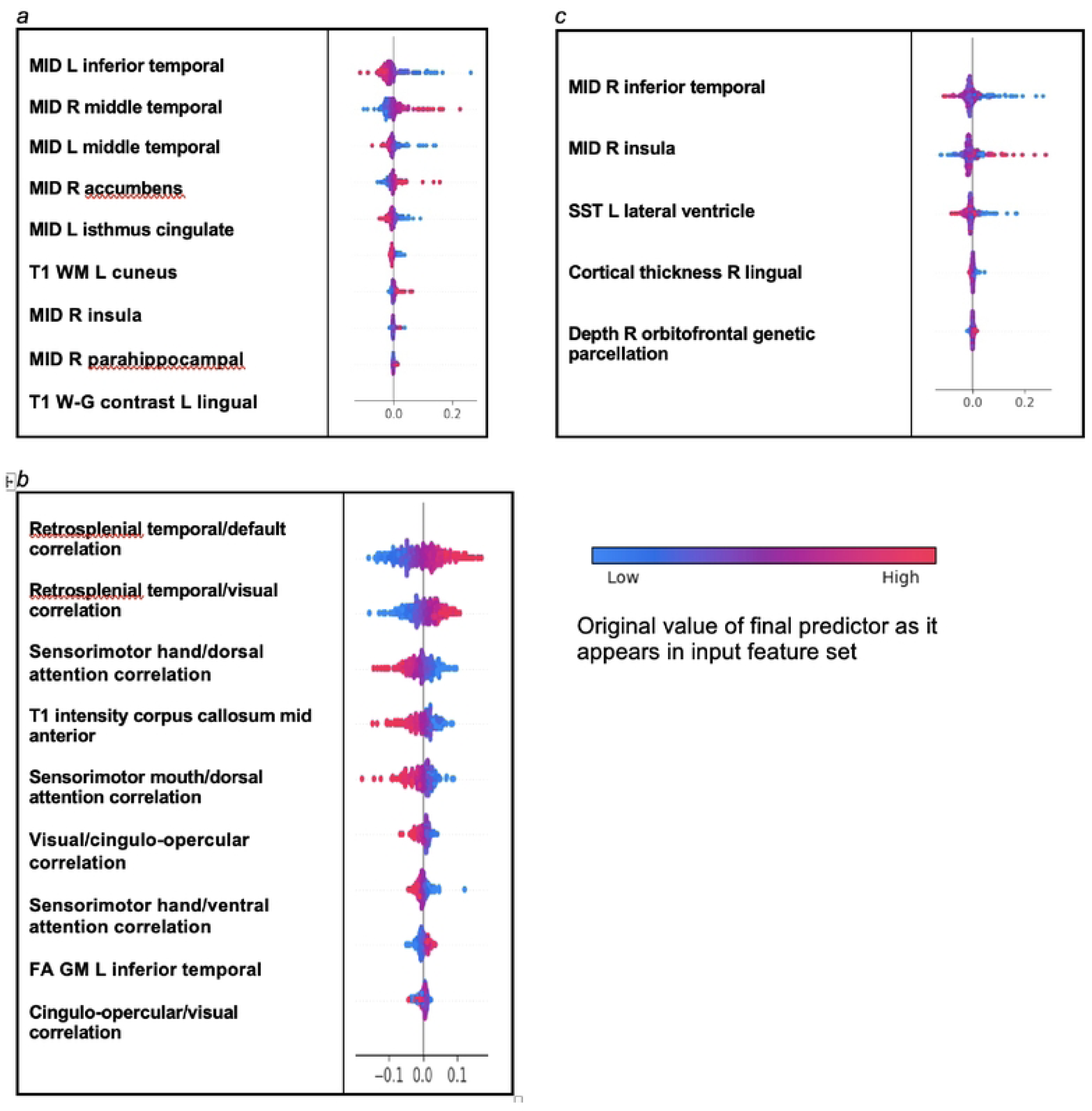
Individual-level importances of neural final predictors of somatic symptom disorder in early adolescence. Summary plots are presented of the importance of each final predictor (computed with the Shapley Additive Explanations technique) on an individual subject level to predicting SSD a) with new onset at 11-12 yrs; b) in all cases at 9-10 yrs; and c) in all cases at 11-12 yrs. The color gradient represents the original value of each feature (metric) where red = high and blue = low. Discrete (binary) features appear as red or blue, while continuous features appear as a color gradient.

### The relationship between accuracy and final predictor importance

We investigated the relationship between accuracy and final predictor importance by computing the mean predictor importance for each experiment. For example, the average importance of final predictors of new onset depression at 11-12 years in testing in held-out, unseen data (**Table 6**). This data may be inspected in **Supplementary Table 3**. We then computed the correlation and R^2^ of the relationship between accuracy and mean predictor importance across all experiments. Across all the experiments described in the present study, the correlation between accuracy and predictor importance in final, optimized models tested in held-out, unseen data was 78.4% and the R^2^ was 61.5%. Interestingly, the two outliers observable in Figure 9 were both multimodal predictive models of Anxiety (new onset cases at 11-12 yrs and contemporaneous prevailing cases at 9-10 yrs), where the very high accuracy of these models placed them off the trendline.

**Figure 9:**
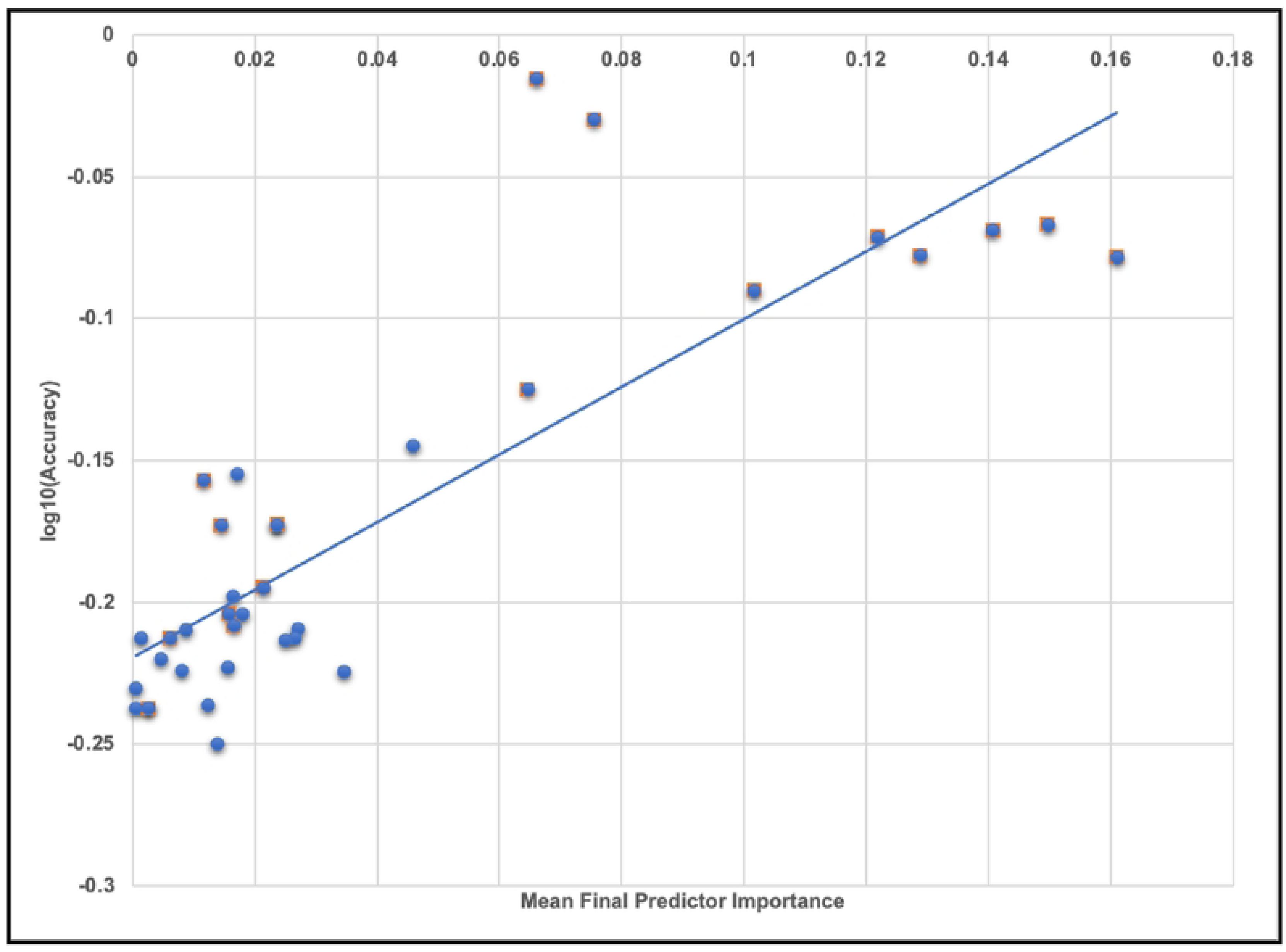
The relationship between accuracy and final predictor importance. Average variable importance computed with the Shapley Additive Explanations technique is shown plotted against the log of prediction accuracy in testing in held-out data for each experiment in the study. The line of best fit obtained with a linear regression is also displayed. Underlying data for this chart may be inspected in Supplementary Table 4.

## DISCUSSION

### Common and specific themes across internalizing disorders

We analyzed ∼6,000 candidate predictors from multiple knowledge domains (cognitive, psychosocial, neural, biological) contributed by children of late elementary school age (9-10 yrs) and their parents and constructed robust, individual-level models predicting the later (11-12 yrs) onset of depression, anxiety and SSD. Leveraging an optimization pipeline that included AI-guided automated feature selection allowed us to extend prior work by analyzing a wider variety of predictor types and ∼40x more candidate predictors than previous comparable ML studies. A common pre-processing and analytic design across all three internalizing disorders in the same youth cohort allows the direct comparison of results to elicit their diagnostic specificity and identify common themes. In addition, we wanted to quantify the relative predictive performance of multimodal vs neural features and examine the relationship between predictor importance and model accuracy. To our knowledge, this is the first ML study in adolescent internalizing disorders to include multiple types of neural predictors (rsfMRI connectivity; task fMRI effects; diffusion and structural metrics), analyze >200 multimodal features and quantify the relationship between predictor importance and accuracy.

Comparing across results, we found that the relative predictive performance of our models varied according to the specific disorder and type of predictor (psychosocial vs neural). Deep learning optimized with IEL rendered robust individual-level predictions of all three internalizing disorders with AUROCs of ∼0.90-0.99% and 81-97% accuracy. Precision and recall were also consistently ≥∼80% with scattered exceptions in precision (new onset depression: 75% and prevailing SSD at 11-12 yrs: 77%). Our primary focus was in predicting future, new onset cases of each internalizing disorder in early adolescence. We found that new onset cases of anxiety could be most reliably predicted (AUROC ∼0.96), followed by depression (AUROC ∼0.90) and SSD (AUROC ∼0.89). A similar differential was also present when predicting contemporaneous prevailing cases at 9-10 yrs but disappeared when considering all prevailing cases at 11-12 yrs. Depression proved a more challenging condition to predict when taken across all experiments, perhaps reflecting its later median age of onset and less-differentiable diagnostic phenotype in adolescence. (46, 47)

Overall, we found that predicting early adolescent internalizing disorders with multimodal features resulted in substantially better performance than exclusively neural-based models, and that psychosocial predictors were preferentially selected in multimodal modeling. Our pipeline includes automated feature selection with a genetic algorithm (IEL) that progressively selects among features as it learns how to optimize predictive models over a principled training process (typically ∼40,000 models). Cognitive, neural and biological features failed to outcompete psychosocial features in training with multimodal features -- with a single notable exception in new onset anxiety. Further targeted experiments specifically assessed the standalone predictive ability of multiple neural feature types derived from MRI. These experiments demonstrated that neural-only models sacrifice 10-25% performance across statistics (accuracy, AUROC, precision, recall) with smaller discrepancies in new onset depression and anxiety, where neural-only features achieved moderately robust 0.77 AUROC and 70% and 75% accuracy, respectively. While little extant research has directly compared psychosocial to neural features in youth internalizing disorders, our results are congruent with studies that have used multimodal feature types including MRI metrics. (18, 19) Our design extended prior work by allowing us to examine more and wider feature types and disorders and the prediction of new onset vs prevailing cases. Neural-only models of new onset cases achieved superior performance to other participant samples and selectively comprised task fMRI and structural metrics, though more neural feature types (rsfMRI connectivity, diffusion-based) were available for selection, suggesting structural and task fMRI neural features may have particular promise in predicting adolescent onset of internalizing disorders.

Specific sets of final predictors for each disorder and participant sample were unique and differentiated both a) depression, anxiety and SSD from each other and b) future new onset from all prevailing cases. However, parental levels of various types of problem behaviors and youth sleep disturbances appeared as cross-cutting, higher-level themes. Depression and anxiety showed closer commonality, with parent externalizing, avoidant and intrusive traits and total problem burden assorting as predictors across different participant samples. Notable disorder-specific predictors included parent level of somaticizing to their child’s SSD and parental anxiety level to their child’s anxiety. Taken together, our results demonstrate that parent problem behavioral traits are important drivers of internalizing disorders in early adolescence and that the specific parental traits observed when their child is 9-10 yrs may be useful in discriminating whether their child will go on to develop depression, anxiety or SSD. This phenomenon suggests intergenerational transmission, though our design cannot determine whether this is underpinned by inheritance, parent-youth styles of relating or other factors, though the presence of externalizing parental behaviors in predicting the later onset of depression and anxiety suggests that more than inheritance is at work. Here, our results congruent with the small number of comparable ML studies that have included parental traits as candidate predictors, where parent total behavioral problems and poor maternal relationships were leading predictors of depression. (15, 48) Sleep disturbances may affect up to ∼40% of elementary school age children and youth with both internalizing and externalizing disorders are at elevated risk. (49, 50) We found that sleep disturbances in the late elementary school age group (9-10yrs) predicted the later (11-12 yrs) onset (anxiety, SSD) and prevalence (depression) of internalizing disorders, congruent with recent research showing that disturbed or short duration sleep predicts later internalizing symptoms. (51, 52, 53, 54) Here, our findings add to a growing body of work suggesting sleep disturbances may be important intervention targets in elementary school age youth to reduce the later burden of internalizing symptoms. (51)

Recent research in association-based studies has suggested that effect sizes in neuroimaging studies of psychopathology and cognitive traits are often inflated, particularly in smaller participant samples, resulting in generalization failure. (55) Accordingly, we investigated predictor importance at both the group and individual level and its relationship with model performance in generalization testing, observing a strong relationship between predictor importance and accuracy across experiments. In individual experiments, psychosocial predictors in multimodal models exhibited larger importances with wider inter-individual importance dispersions than those in neural-only experiments, even after extensive optimization and principled feature selection. Collectively, these results suggest that the smaller importances of neural features - and perhaps their more restricted variability among individuals - were at least related to their weaker performance in predicting cases using artificial neural networks. Future work will be required to determine whether these phenomena are seen in other disorders and participant samples (particularly other developmental periods) and if other types of neural features (for example, connectivity features obtained from data-driven rather than ROI methods) could fare better in predicting cases of internalizing disorders.

### Depression

Depression is a common and growing problem in adolescence which elevates later risk for suicide, poor educational outcomes and substance use. (56) In the present study, we focus on early onset cases of depression i.e. those which onset or are present at 11-12 yrs. Most prior work in early onset depression has examined psychosocial predictors at the group level, linking it to sleep disturbances, childhood adverse events (neglect, abuse, loss of parent), familial depression and pubertal changes (57, 58, 59, 60, 61, 62, 63) Longitudinal neuroimaging studies of the onset or course of depression in adolescence are relatively plentiful and have ranged across a variety of MRI modalities. (64) Similarly, these have typically been group-level studies employing traditional multivariate predictive methods in a single neuroimaging modality and small number of ROIs, sometimes in small samples. Results have been inconsistent. In structural MRI, subcortical regions (especially hippocampal) have been most intensively studied with mostly negative results, though there is some evidence for smaller accumbens and insula volume and equivocal results for OFC regions. (65, 66, 67, 68, 69, 70) In fMRI, reward and emotion processing have been most intensively studied. A number of studies have demonstrated differential reward-related activity in the ventral striatum, (71, 72, 73, 74, 75) though these studies are nearly all from later adolescence. In early adolescence, Morgan et al found the inverse was the case. (76) In emotion processing, increases or decreases in ACC activity have predicted adolescent depression onset. (77, 78, 79)

More recently, a number of ML studies have performed prospective prediction of adolescent depression incorporating larger numbers of candidate predictors, either psychosocial and/or neuroimaging. To our knowledge, our study represents only the second time multimodal (including neuroimaging) candidate predictors have been analyzed at the individual level using ML to prospectively predict depression onset in adolescents, and the first time in early adolescence. With an AUC of ∼0.90, we achieved performance comparable with a single prior deep learning study and superior to that obtained using logistic regression or support vector machines (SVM). (18, 48, 70, 80, 81) We are not aware of other prior ML studies that have directly compared the ability of multimodal vs neuroimaging predictors in adolescent depression or incorporated more than one type of neuroimaging metric.

Our AI-guided optimization pipeline preferentially selected psychosocial features to predict early onset adolescent depression after analyzing thousands of multimodal candidate predictors. Multimodal models achieved 10-15% better performance over all metrics than neural-only models. However, at ∼0.77 AUROC, our neural-only deep learning model achieved performance similar or better to multimodal models in other studies using different ML methods (logistic regression, SVM). Several recent large-scale ML prospective predictive studies of youth depression have examined the predictive performance of nonlinear combinations of candidate predictors at the individual level. In youth aged 15 yrs, Rocha et al trained penalized logistic regression models with 11 psychosocial metrics finding that school failure, social isolation, involvement in physical fights, drug use, running away from home, and maltreatment predicted depression onset at 18 yrs, achieving AUROC 0.79 in the baseline dataset and 0.59 and 0.63 in external validation datasets. Foland-Ross et al used cortical thickness metrics to predict new onset adolescent depression with 70% accuracy, with thickness of the right precentral and medial OFC and left ACC and insula representing the most important features. Most recently, two important large scale ML studies utilized multimodal candidate predictor sets. Toenders et al applied penalized logistic regression to 69 phenotypic and 76 structural MRI metrics in youth aged 14 yrs from the IMAGEN dataset, testing for generalization in a held-out set to achieve 0.72 AUROC and 66% accuracy. Depressive symptoms at baseline, neuroticism, cognition, supramarginal gyrus surface area, and stressful life events were most predictive of later new onset depression. Xiang et al surveyed 188 psychosocial and rsfMRI connectivity candidate predictors collected at 9-10 yrs and empirically selected based on prior literature to predict depression trajectories (computed with latent class analysis) through 11-12 yrs in the ABCD cohort, with deep learning achieving best performance. This study is perhaps the most comparable to our own methodologically and achieved similar AUROC (∼0.90) and accuracy (87% vs ∼82, ∼86%), though precision (0.45) and recall (0.44) were lower. Total sleep disturbance, parent total behavioral problems, financial adversity, ventral attention-left caudate and dorsal attention-left putamen connectivity and school disengagement were the most important predictors of depression trajectories. Thus, we obtained thematically concordant results with prior research in identifying parental problem behaviors of various types and sleep disturbances being important predictors of early adolescent depression. However, our work differs in not identifying other types of childhood adverse experiences, cognitive traits and pubertal status as being as important to final, optimized models. In new onset depression, we found that parent externalizing behaviors were the most important predictor followed by whether the child had come to clinical attention prior to 9-10 yrs and inverse relationships with secondary caregiver acceptance and degree of prosocial behaviors. In contrast, parent avoidant and intrusive behaviors and sleep disorder of excessive somnolence drove the prediction of all prevailing cases at 11-12 yrs.

We believe that this is the first time that multiple neuroimaging feature types have been used to predict new onset depression in adolescence in a neural-only model. Thus, it is particularly intriguing to note that the onset of early adolescent depression was predicted by multiple task fMRI effects – but that these centered on the SST (which measures response inhibition) rather than the MID (reward processing). We found rather that MID effects were emphasized in predicting anxiety and SSD -- and it has been previously noted that almost no longitudinal fMRI studies in adolescent depression directly compare anxiety and depression in the same sample. (64) In our neural-only models, early onset depression was predicted by SST effects in the left caudal anterior cingulate, pars opercularis, entorhinal and right parstriangularis (inverse relationships) and left transverse temporal and accumbens (positive directionality) ROIs. As well, structural gray matter features in the right OFC and precentral ROIs were important predictors. Thus, our results are concordant with existing literature in highlighting OFC and accumbens ROIs but our algorithms preferentially selected effects from the SST over the MID. The SST is a test of inhibition of prepotent responses and has been extensively studied in externalizing disorders (where there is a positive relationship) but less in the internalizing disorders, where we identified it has a negative relationship with depression. However, ex-scanner studies in children with internalizing behaviors and adults with depression using the SST show longer reaction time in patients with recent work associating response inhibition deficits in children with rumination traits. (82, 83, 84) Future work may consider exploring SST task-related effects in response inhibition further in adolescent depression. Lastly, we note that metrics of head motion in the SST appeared as final predictors in depression. Because our intent was to perform large-scale data-driven ML prediction and all MRI metrics had passed quality control, we treated head motion metrics *pari passu* with other feature types and did not exclude participants based on head motion thresholds (as is commonly done in specialist neuroimaging studies). While head motion metrics were included in all analyses, they only appeared as final predictors in depression. While this could be considered a nuisance, we also note that response inhibition in the SST has been previously associated with bursts of antagonistic neck muscle activity due to a compensatory vestibular-ocular reflex consistent with the saccadic race model, and the latter may be worth investigating further in the context of adolescent depression. (85, 86)

### Anxiety

Anxiety is among the most common mental health disorders affecting adolescents and adults. Among the internalizing disorders, it is the condition most clearly centered on early adolescence, with a median age of onset of 11 yrs. Many psychosocial, demographic and cognitive risk factors have been associated with the development of clinical anxiety including early life temperamental traits such as anxiety sensitivity, neuroticism and anxious temperament. Thus, the formulation of prospective predictive models that can discriminate among these factors and provide reliable, individual-level predictions of anxiety onset in early adolescence is of particular relevance. However, few ML studies have predicted future anxiety in adolescence. To our knowledge, this is the first ML study to predict future anxiety in early adolescence and the first to use multiple neural features types. In important prior multimodal work, Chavanne et al compared the ability of psychosocial vs neural features to predicting anxiety cases at 18-23 yrs in the IMAGEN cohort with 14 gray matter volumetric measures and 13 clinical metrics measured at 14 yrs using a majority voting algorithm comprising Logistic Regression, SVM and Random Forest classifiers. In the multimodal model, an AUROC of 0.68 was obtained with neuroticism, hopelessness, emotional symptoms and family factors contributing most to the prediction and volumetric differences in the periaqueductal gray, amygdala, ACC and subcortical regions making lesser contributions. With neural features alone, AUROC dropped to 0.52 whereas with psychosocial features alone it improved to 0.69.

Here, we demonstrate that new cases of anxiety at 11-12 yrs can be very reliably (AUROC ∼96%; accuracy and precision ∼93%) predicted with deep learning optimized with IEL and that these predictive models differ from depression and SSD. As in the developmentally older IMAGEN cohort, our analysis in the younger ABCD cohort found that multimodal features predict the onset of anxiety better than neural-only features with a substantial differential of 15-20% across performance statistics. However, the neural-only model achieved respectable performance in the context of the literature as a whole, with AUROC of ∼78% and accuracy of 75%. We found that new onset cases of anxiety in early adolescence were predicted by the child having received clinical services prior to 9-10 yrs, total sleep disturbance, mother’s mental health clinical history and parent levels of problem externalizing and avoidant behaviors. There were inverse relationships with MID large vs small loss contrast in the left OFC, whether parent and youth got along well, and prosocial behaviors. This is a particularly interesting result since it is the only one among our experiments where a neural feature ‘outcompeted’ thousands of other candidate predictors in the AI-guided optimization process to survive into the final, optimized model. While the MID is perhaps best known for measuring reward seeking, it also measures the avoidance of punishment and loss anticipation. Activations in ventro-lateral prefrontal regions, median cingulate cortex and the amygdala are specific to loss events. (87) The ventro-lateral prefrontal cortex and OFC are localized sub-regions of the ventral prefrontal cortex that both underpin social flexibility, with the OFC being well-associated with responses to the positive and negative valence of social stimuli. (88) Structural OFC changes have been associated with adolescents who experienced negative interactions with their mother. (89) Our results are congruent with this literature and suggest that aversive parent-child factors (externalizing and avoidant styles, tenor of the parent-child relationship, poor maternal mental health), sleep disturbances, social withdrawal and pre-existing differences in the anticipation of loss in the OFC in childhood (before or at ages 9-10 yrs) interact in a nonlinear manner to predict the onset of later clinical anxiety in early adolescence. While there was thematic overlap among our different anxiety models (parent problem behaviors, sleep disturbances, social/peer relationship factors) this particular set of factors was specific to new onset cases. While parent anxiety problem behaviors were a final predictor of contemporaneous cases at 9-10 yrs, they did not predict new onset cases at 11-12 yrs. Similarly, no neural predictors appeared in the multimodal models predicting prevailing cases at either 9-10 yrs or 11-12 yrs.

### Somatic Symptom Disorder

Somatic behavioral problems refer to the presence of one or more physical symptoms accompanied by excessive investment (time, emotion, behaviors) in the symptom(s) that results in significant distress or dysfunction. The diagnosis of SSD emphasizes symptom-based impairment in daily life. Peri-adolescence is an important period when SSD onsets and rises towards higher adult rates. Prior research, including prospective studies, has frequently implicated family functioning including parents’ own levels of physical and mental health complaints and parent somatic problems as well as parental divorce, illness or death, childhood traumatic experiences and insecure attachment. (90, 91, 92, 93, 94, 95) Work examining adolescent predictors of subsequent trajectories of somatic symptoms have identified the quality of parent-youth relationships, parenting stress and youth bullying, school dissatisfaction and lower intelligence level symptoms as important predictors. (96, 97, 98, 99, 100) The genetic component appears to be small, albeit studies are limited. (101) Research focused on the cognitive-affective neural basis of somatic problems using task fMRI has linked group-level differences in para/hippocampal, ACC, insula, brainstem and lateral prefrontal regions to effects in negative expectancy, attentional bias and pain catastrophizing. (102, 103, 104, 105, 106, 107, 108) Fewer neuroimaging studies have investigated circuit abnormalities in somatic problems, though rsfMRI studies have implicated increased brainstem, caudate, thalamus and ACC activity and decreased lateral prefrontal activity in adults. (109, 110) In a cross-sectional study in the ABCD cohort, Dhamala et al found disrupted temporo-parietal, default mode, dorsal attention and control-limbic functional connections using rsfMRI data from 9-10 yrs to predict CBCL somatic problem scores at the same age. (111)

Our findings contribute to this growing body of work in several ways. Firstly, prospective predictive studies of somatic problems have typically focused on either psychosocial (particularly family- or adversity-related measures) or neural predictors. In the present study we analyzed nearly 6,000 multimodal predictors of many types (including cognitive and non-neural biological metrics), allowing us to assess their relative predictive ability holistically. In these multimodal models, we found that psychosocial predictors were preferred over neural, cognitive and biological metrics. Secondly, the richness of parent and family-related metrics in the ABCD sample allowed us to consider a larger range of psychosocial predictors than has typically been available to earlier studies of somatic problem symptoms in youth. We found that parent level of somatic problem behaviors (new onset cases, 9-10 yr prevailing cases) and internalizing traits (11-12 yr prevailing cases) were preferentially selected as predictors over other family-, school- or peer-related candidate predictors such as bullying, parent stress or early adverse experiences. In all participant samples, parent somatic or internalizing problem behaviors interacted with sleep disturbances. Of note, whether a youth had come to clinical attention for a mental health issue predicted the later onset of somatic problems and a specific predictor of somatic problems in cases at 9-10 yrs was whether the child was seen for a medical issue other than a regular checkup. These findings comport with earlier work and further suggest that childhood patterns of clinical use and sleep disturbances and elevated levels of parent somatic traits may be helpful in assessing youth risk for somatic problem behaviors. Similarly, the wide range of neuroimaging measures available allowed us to assess nearly 5,000 different neuroimaging metrics over multiple modalities to predict somatic problem behaviors in youth. While these models were not as robust as multimodal models (AUROC ∼0.64-0.70), they are congruent with extant research in centering on parahippocampal, temporo-parietal, cingulate ROIs and default mode and attentional network connectivity. Our work additionally highlighted the insula, a region long known to be involved in interoception and pain processing. Interestingly, effects in these regions during the MID task involving reward processing and loss anticipation were emphasized in predicting new onset cases of somatic problems in contrast to anxiety, where they centered on loss anticipation only. While we are not aware of prior work using the MID task in somatic problem behaviors, this may be an interesting line of future inquiry given a cardinal feature of somatization is the amount of valence and/or investment given to physical symptoms. Overall, we found that structural, task and rsfMRI were useful modalities in predicting somatic problems in early adolescence but diffusion imaging made less of a contribution.

## LIMITATIONS

This study uses secondary data from the ABCD study and we were therefore unable to control for any bias during data collection. While the ABCD study strived for population representation, there is a mild bias toward higher-income participant families of white race in the early adolescent cohort. Data is not available prior to baseline (age 9-10 years) assessment and we cannot conclusively rule out that youth participants met criteria for depression, anxiety or somatic problems prior to this age but not at baseline assessment at 9-10 years of age. Thus, it is possible that certain cases coded as ‘new onset’ at 11-12 years of age in our analysis could have met clinical criteria ≤8 yrs but were in remission at 9-10 yrs. In the present study, we defined cases as any individual meeting ASEBA clinical thresholds in the CBCL subscale scores of interest and did not exclude participants who thereby met criteria for other conditions. Thus, co-morbidity may be present in the experimental samples as is common in clinical populations and in most research studies in early adolescence. While we used nearly 6,000 variables available in the ABCD dataset, our study is not exhaustive. It is possible that different results could have been obtained if more or different candidate predictors were included. For example, rsfMRI data includes metrics from ROI-based parcellations but not a data-driven method such as ICA. We tested for generalization in a holdout, unseen test set obtained by partitioning the data, a gold standard method in ML. However, methods and results should also be tested for replication in an external dataset other than ABCD.

## Data Availability

Data used in the preparation of this article were obtained from the Adolescent Brain Cognitive Development (ABCD) Study (https://abcdstudy.org), held in the NIMH Data Archive (NDA). Details regarding how to obtain this data are provided in the Acknowledgements section of the manuscript as recommended by ABCD and NIMH. All relevant data created in this study are within the manuscript and its Supporting Information files.

https://nda.nih.gov/abcd/abcd-annual-releases.html

## ACKNOWLEDGEMENTS

Research reported in this publication was supported by the National Institute of Mental Health of the National Institutes of Health under award number **R00MH118359** to NdL. The content is solely the responsibility of the authors and does not necessarily represent the official views of the National Institutes of Health. The support and resources from the Center for High Performance Computing at the University of Utah are also gratefully acknowledged.

Data used in the preparation of this article were obtained from the Adolescent Brain Cognitive Development^SM^ (ABCD) Study (https://abcdstudy.org), held in the NIMH Data Archive (NDA). This is a multisite, longitudinal study designed to recruit more than 10,000 children age 9-10 and follow them over 10 years into early adulthood. The ABCD Study® is supported by the National Institutes of Health and additional federal partners under award numbers U01DA041048, U01DA050989, U01DA051016, U01DA041022, U01DA051018, U01DA051037, U01DA050987, U01DA041174, U01DA041106, U01DA041117, U01DA041028, U01DA041134, U01DA050988, U01DA051039, U01DA041156, U01DA041025, U01DA041120, U01DA051038, U01DA041148, U01DA041093, U01DA041089, U24DA041123, U24DA041147. Additional support for this work was made possible from NIEHS R01-ES032295 and R01-ES031074. A full list of supporters is available at https://abcdstudy.org/federal-partners.html. A listing of participating sites and a complete listing of the study investigators can be found at https://abcdstudy.org/consortium_members/. ABCD consortium investigators designed and implemented the study and/or provided data but did not necessarily participate in the analysis or writing of this report. This manuscript reflects the views of the authors and may not reflect the opinions or views of the NIH or ABCD consortium investigators. The ABCD data repository grows and changes over time. The ABCD data used in this report came from <u>10.15154/1523041</u>. DOIs can be found at https://nda.nih.gov/abcd/abcd-annual-releases.html.

## Notes

### Competing Interest Statement

The authors have declared no competing interest.

### Author Declarations

University of Utah Institutional Review Board

